# Longitudinal Drain-Fluid Proteomics Reveals Procedure-Specific Signatures of Postoperative Leakage after Pancreatic and Colorectal Surgery

**DOI:** 10.64898/2026.09.17.26363291

**Authors:** Alexander Jessernig, Victoria M. Agboola, Sibylle Pfammatter, Ignazio Tarantino, Robert Polak, Vaclav Liska, Inge K. Herrmann

## Abstract

Anastomotic leakage is a major complication after gastrointestinal surgery, yet early detection remains limited by nonspecific clinical signs and narrow biomarker panels. Here, we used longitudinal data-independent acquisition proteomics to profile postoperative drain fluid from two surgical cohorts, comprising 33 pancreatic surgery patients with 201 drain-fluid samples and 34 colorectal surgery patients with 143 drain-fluid samples with an observation window of up to 10 days postoperatively. We aimed to define procedure-specific leakage signatures, evaluate established biomarkers across postoperative time, and identify additional candidate markers beyond routinely measured proteins. In pancreatic surgery, established enzyme markers, particularly AMY2A and PNLIP, showed strong early discrimination between leak and non-leak patients, while in colorectal surgery, the same predefined marker strategy performed less consistently. Discovery analysis revealed divergent temporal patterns. Pancreatic leakage was dominated by early (post-operative day (POD) 1-3) enzyme enrichment followed by inflammatory, antimicrobial, oxidative stress, and matrix-remodeling-associated proteins. Colorectal leakage showed strongest divergence around POD3 and was driven largely by reduced abundance of proteins linked to wound healing, including endothelial integrity, extracellular matrix organization, complement regulation, and coagulation balance. From these procedure-specific patterns, PRSS2 and AZU1 were found for pancreatic surgery, and FBLN1 and CDH5 for colorectal surgery, as novel candidate markers for future follow up validation studies establishing their diagnostic potential. Overall, these data show that postoperative leakage follows procedure-specific proteomic trajectories and support drain-fluid proteomics as a framework for biologically informed biomarker discovery beyond routinely measured markers.

## INTRODUCTION

Anastomotic leakage (AL) remains one of the most dreaded complications after gastrointestinal surgery. Despite advances in surgical technique and perioperative care, failure of an anastomosis continues to increase postoperative morbidity and mortality.^1,2^ Early detection is key to improving patient outcome. Yet, in clinical practice, leakage is frequently recognized only after local failure has progressed to systemic deterioration. Clinical signs such as fever, abdominal pain, tachycardia, ileus, and leukocytosis overlap with the physiological inflammatory response to surgery, while computed tomography is typically performed only after clinical suspicion has already emerged.^3–5^ This creates a clinically important diagnostic window in which biologically active leakage may already be present but remains difficult to detect. The disconnect between early local pathology and clinical recognition has made biomarker-based monitoring an attractive strategy for postoperative surveillance. To sample this window, clinical practice already provides a direct route to the anastomotic site. Prophylactic drains are routinely placed adjacent to the anastomosis at the end of gastrointestinal surgery, both to remove perianastomotic collections and to permit early identification of a developing leak.^6^ The fluid recovered from these drains is continuously and non-invasively accessible at the bedside. Rather than an inert surgical by-product, drain fluid is a dynamic source of information shaped by perianastomotic pathophysiology, whose composition reflects the local state of the anastomosis. Additionally, when intraluminal contents escape through a defect, they carry a distinct biochemical signature into this otherwise sterile compartment. These properties make drain fluid an appealing matrix for postoperative surveillance, although drain type, position relative to the anastomosis, as well as the timing of sampling and removal can introduce variability in the quality of the information that can be obtained.

The biomarkers that are being analyzed from drain fluid differ markedly between surgical settings. After pancreatic resection, drain fluid amylase is the dominant analyte and is embedded in the consensus definition of postoperative pancreatic fistula, defined by the International Study Group as a drain amylase concentration greater than three times the upper limit of the normal serum value on or after the third postoperative day and graded by clinical severity.^7^ Measured as early as the first postoperative day, drain amylase estimates the risk of clinically relevant fistula, although reported thresholds vary widely across cohorts, from a few hundred to several thousand units per liter, mainly caused by differences in populations and derivation methods rather than a stable operating point.^6,8,9^ In colorectal surgery the evidence is more heterogeneous. C-reactive protein, studied most extensively in blood serum or drain fluid, is complemented by drain fluid amylase, which rises sharply in the presence of an early leak,^10,11^ and by peritoneal inflammatory and ischemic markers such as interleukin-6, tumor necrosis factor-α, and the lactate-to-pyruvate ratio.^12^

Despite the biological complexity of anastomotic failure, biomarker selection for AL detection has remained remarkably narrow (Figure 1a). Across gastrointestinal surgery, and irrespective of the differing tissue and enzymatic environment of each anastomosis, leak detection converges on the same limited selection of acute-phase and cellular inflammatory markers, ^13–16^, digestive enzymes, ^17–21^ and selected cytokines..^12,22^ This reductive approach contrasts with the molecular complexity of postoperative drain fluid. Its protein composition may therefore capture early changes at the anastomotic site that are not accessible with routinely measured markers.

**Figure 1:**
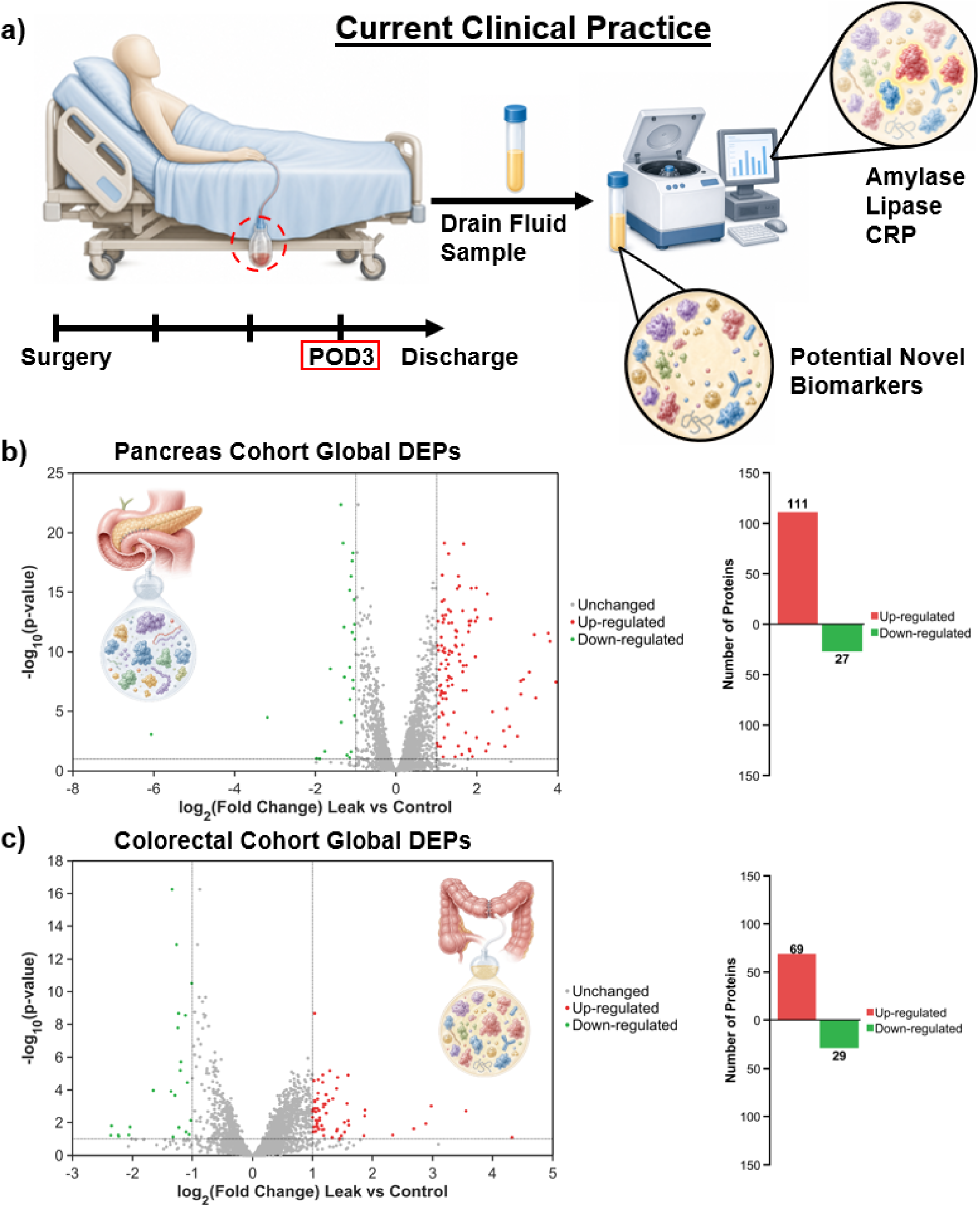
Global differential expression across postoperative days in pancreatic and colorectal cohorts. a) Depiction of current clinical practice. Only a small fraction of biomarkers is used for AL detection while the drain fluid contains a complex mixture of potentially novel biomarkers. b) Volcano plot showing differential protein abundance between leak and non-leak patients in the pancreatic cohort based on all POD pooled data. Proteins are plotted by log2 fold change and −log10 p value. Red indicates proteins increased in leak, green indicates proteins increased in non-leak, and gray indicates non-significant proteins. Bar plot summarizes the number of significantly increased proteins in each group (111 in leak, 27 in non-leak). c) Volcano plot showing differential protein abundance in the colorectal cohort under the same criteria. Compared to the pancreatic cohort, fewer proteins reach large effect sizes and significance. Bar plot shows the number of significantly increased proteins (69 in leak, 29 in non-leak).

Mass spectrometry-based proteomics is well suited to access the broader drain-fluid biology, as proteomic profiling does not require prior selection of individual analytes, but enables parallel quantification of large parts of the protein landscape present at the surgical site. In rectal cancer surgery, preliminary proteomic profiling of postoperative drain fluid has identified 1071 proteins and 248 differentially expressed proteins across two POD’s, some of which had not previously been explored as leakage biomarkers.^23^ However, this study was limited to a single surgical setting and sampled only two postoperative time points, leaving the temporal and procedure-specific structure of drain-fluid leakage biology unresolved.

Here, we performed longitudinal proteomic profiling of postoperative drain fluid from pancreatic and colorectal surgery cohorts obtained from two centers to define molecular signatures associated with leakage across two distinct gastrointestinal surgical settings. We first evaluated previously proposed leakage biomarkers within the proteomic dataset to assess their reproducibility across procedures and postoperative time points. We then applied unbiased discovery analysis to identify additional proteins and biological pathways associated with leak development. This approach directly compares established biomarker paradigms with the broader molecular information contained in drain fluid and provides a framework for procedure-specific biomarker discovery in postoperative leak detection.

## RESULTS AND DISCUSSION

### Sample Availability and Data Stability

We first sought to determine whether postoperative drain fluid is associated with distinct proteomic signatures in pancreatic and colorectal surgery. Global differential abundance analysis between leak and non-leak patients revealed clear surgical procedure-specific differences in both the magnitude and directionality of proteomic divergence (see Figure 1b,c). In the pancreatic cohort, the volcano plot showed a pronounced leak-associated shift, with 111 proteins enriched in leak samples and 27 proteins enriched in non-leak samples. Many of these proteins displayed large positive log2 fold changes and low FDR values. By contrast, the colorectal cohort showed a more moderate global separation, with fewer proteins meeting the differential abundance criteria and smaller fold changes. Across postoperative days, there was generally little overlap in significantly elevated proteins between the pancreatic and colorectal cohorts (see Figure 2a). The greatest cross-cohort overlap was observed on POD3, with many of the shared proteins associated with inflammatory and innate immune response in the leak group. These results indicate that leakage-associated proteomic changes are detectable in both cohorts, but that the pancreatic cohort is characterized by a stronger and more directional global response (for POD specific Vulcano plots see Figure S1 and S2 in the SI).

**Figure 2:**
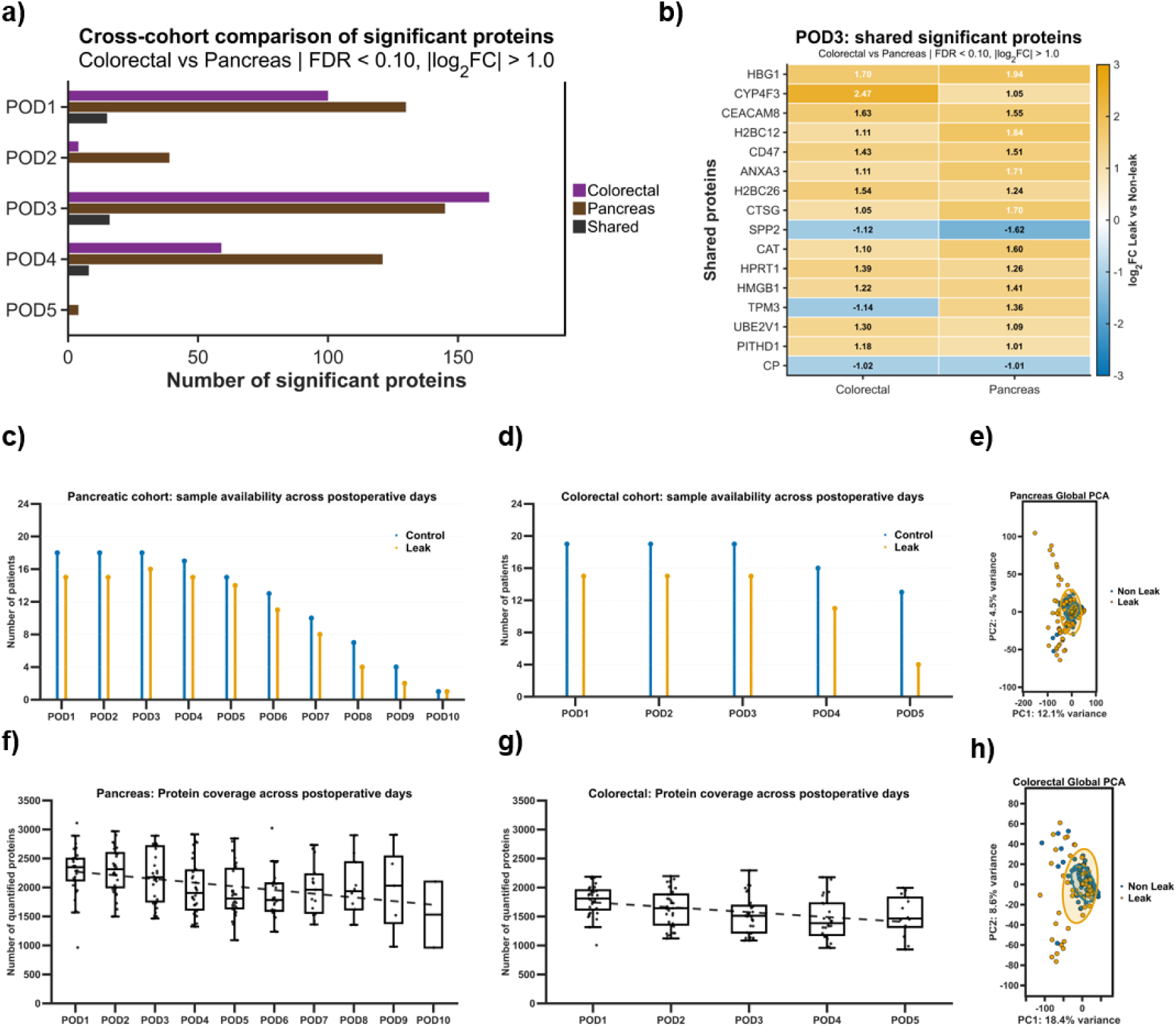
Overview of cohort sample availability and protein coverage. a) Cross-cohort comparison of significant proteins. Most significant proteins were not shared between surgical procedure with a peak on POD3 b) Overview of most significant shared proteins on POD3. A clear trend toward inflammation and oxidative stress is visible in the leak patients. c) Patient availability across postoperative days for the pancreatic cohort. Bar plots show the number of non-leak (blue) and leak (orange) patients per postoperative day. The pancreatic cohort spans POD1 to POD10 with decreasing sample numbers at later days. d) Patient availability across postoperative days for the colorectal cohort. The colorectal cohort includes POD1 to POD5 with stable early group sizes and reduced availability at later time points. f,g) Distribution of quantified proteins per sample across postoperative days in the pancreatic cohort f) and colorectal cohort g). Boxplots show median and interquartile range with whiskers indicating data spread. e,h) PCA analysis of the global dataset over all PODs of the pancreatic e) and the colorectal cohort h).

To support interpretation of these global differences, we next assessed patient availability and proteome coverage across postoperative days to determine whether procedure-specific proteomic patterns were supported by comparable longitudinal sampling. Early postoperative days contained comparable numbers of leak and non-leak patients in both cohorts, providing a robust basis for direct temporal comparison from POD1 onward (see Figure 2c,d). The pancreatic cohort extended from POD1 to POD10, allowing assessment of a longer postoperative trajectory, although sample availability declined after POD5 and became limited at later time points, particularly among leak patients. The colorectal cohort covered POD1 to POD5, with balanced group sizes through POD3 and decreasing leak sample availability thereafter. Thus, both cohorts support comparison of early leakage-associated proteomic evolution, whereas later time points require cautious interpretation because of reduced sample numbers.

Proteome coverage was broadly maintained across postoperative days in both cohorts (see Figure 2f,g). Although the number of quantified proteins showed a modest downward trend over time, especially in the pancreatic cohort, distributions remained sufficiently overlapping to support comparability between samples.

Together, the longitudinal sampling structure and broadly maintained proteome coverage provide a suitable basis for interpreting postoperative day-specific changes in leak-associated protein abundance.

Furthermore, principal component analysis was used to assess the global structure and temporal stability of the drain-fluid proteome in both surgical cohorts. In the pancreatic cohort, the global PCA (Figure 2e) showed substantial overlap between leak and non-leak samples, with PC1 and PC2 explaining 12.1% and 4.5% of total variance, respectively. This indicates that leakage-associated differences affect selected components of the proteome rather than causing complete separation of the global proteomic profile. However, leak samples were more broadly distributed, particularly along PC2, suggesting greater interpatient heterogeneity among patients with leakage. POD-specific PCA confirmed this pattern across POD1 to POD8 (see Figure S3a in the SI), with partial overlap between groups at all time points but consistently broader dispersion among leak samples, especially from POD4 onward. In the colorectal cohort, the global PCA (see Figure 2h) similarly showed substantial overlap between leak and non-leak samples, with PC1 and PC2 explaining 18.4% and 8.6% of total variance. Leak samples were more widely distributed than non-leak samples, suggesting greater variability in the proteomic response among patients who developed leakage. Across POD1 to POD5 (see Figure S3b in the SI), the strongest temporal difference was observed on POD3, where non-leak samples formed a compact central cluster while leak samples were more widely distributed. Thus, PCA revealed a shared overall pattern in both cohorts, where leakage was associated with increased proteomic heterogeneity but not complete global separation. However, the temporal behavior differed between surgery types. In the pancreatic cohort, heterogeneity was sustained across multiple postoperative days and became more pronounced from POD4 onward, consistent with an ongoing and variable response to pancreatic leakage. In contrast, the colorectal cohort showed its clearest divergence during a narrower postoperative window, with the strongest separation on POD3. Together, the PCA results support overall sample comparability and show that leakage-related proteomic changes are variable between patients and differ between pancreatic and colorectal surgery.

### Validation of Clinically Established Biomarkers

We next evaluated clinically established leakage-associated markers across postoperative days and surgical cohorts to determine whether routinely investigated enzymes and inflammatory proteins differ between leak and non-leak patients. Proteins were selected based on prior reports and assessed using temporal log2 fold-change trajectories, patient-level abundance distributions, and ROC-based diagnostic performance.

In the pancreatic cohort, digestive enzymes showed strong and early separation between leak and non-leak patients. AMY2A displayed the largest effect sizes, with log2 fold changes of 4.5 at POD1 and 5.0 at POD2 (see Figure 3a) and remained elevated across the postoperative course. Patient-level distributions showed near-complete separation during the earliest postoperative days (Figure 3c), and ROC analysis confirmed excellent diagnostic performance, with AUC values of 1.00 on POD1 and 0.98 on POD2 (Figure 3b). These values exceeded commonly reported ranges for drain amylase after pancreatic surgery, which typically lie around 0.80 to 0.92.^20,24–26^ PNLIP showed a similar pattern, with consistent elevation in leak samples from POD1 onward and fold changes generally between 2 and 4 across early postoperative days (Figure 3c). This translated into strong early discrimination, with an AUC of 0.96 on POD1 (Figure 3b), consistent with previously reported lipase-based discrimination after pancreatic surgery.^24,27–30^ Together, these findings show that pancreatic enzyme markers retain strong diagnostic value, consistent with the biology of pancreatic fistula formation, where disruption of the pancreatic duct exposes the surgical site to enzyme-rich fluid.^25,27,28^

**Figure 3:**
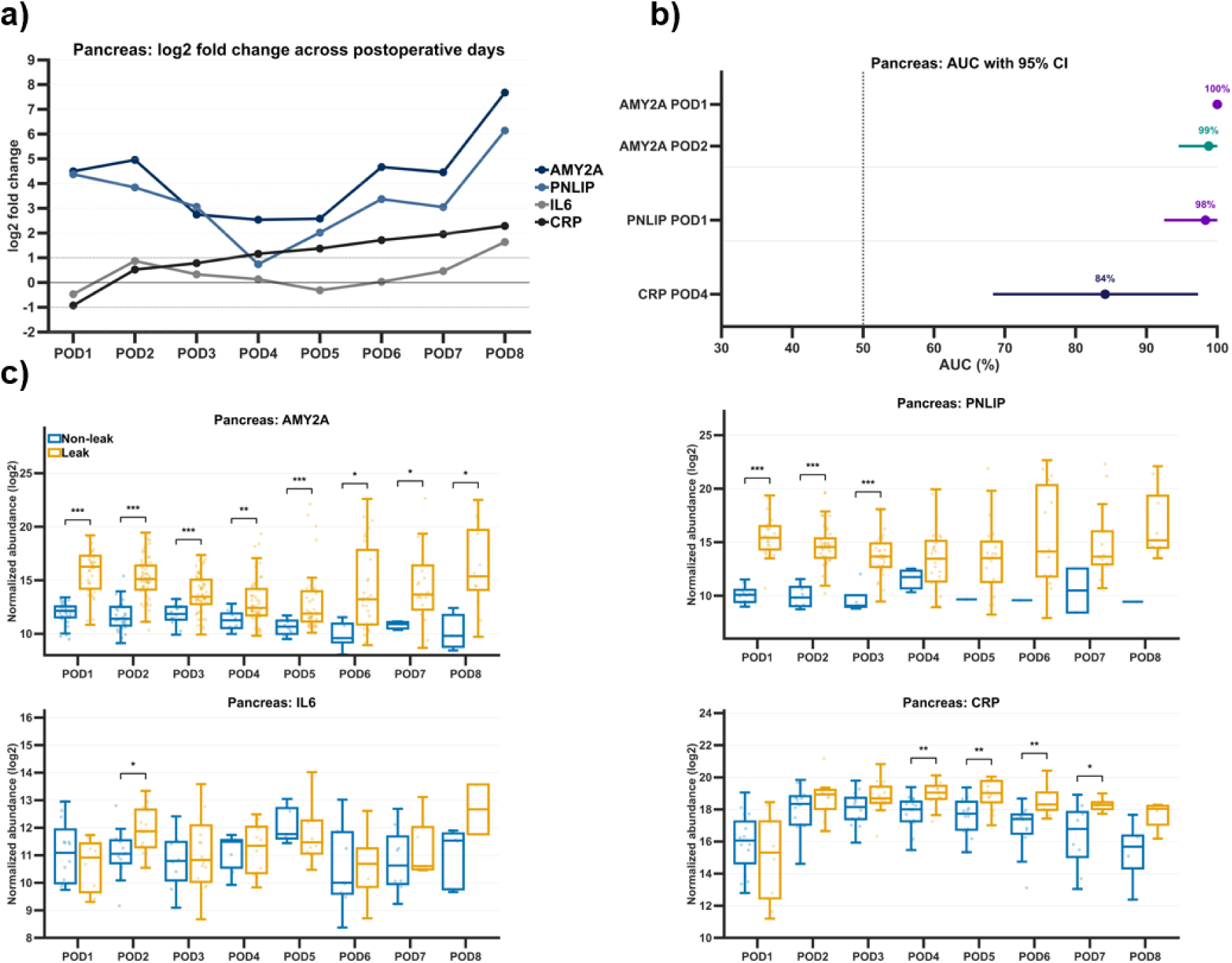
Temporal behavior and patient level distributions established biomarkers markers in the pancreatic cohort. a) Log2 fold changes between leak and non-leak samples across postoperative days for AMY2A, PNLIP, CRP, and IL6. Positive values indicate higher abundance in leak samples. b) Receiver operating characteristic curves for established biomarkers. c) Distributions of normalized protein abundance for individual markers across postoperative days, shown separately for non-leak and leak samples. AMY2A and PNLIP show clear separation at early postoperative days with minimal overlap. CRP shows delayed increase over time. IL6 shows no consistent separation across postoperative days. (* p<0.05, ** p<0.01, ***p<0.001)

Inflammatory markers in the pancreatic cohort showed a different temporal pattern. CRP increased more gradually and achieved its clearest separation at later postoperative time points (consistent with reported timing^25,31^, Figure 3c), reaching an AUC of 0.84 on POD4 and 0.84 on POD5 (Figure 3b). This delayed CRP increase suggests that CRP captures a later inflammatory component of the postoperative response rather than the initial pancreatic enzyme release reported by AMY2A and PNLIP. IL6 showed limited and inconsistent separation, with broad overlap between leak and non-leak patients (Figure 3c). Although IL6 reached moderate discrimination at POD2 with an AUC of 0.77 (consistent with reporting,^32^ Figure 3b), its overall temporal pattern was not stable enough to support strong standalone diagnostic value in pancreatic drain fluid.

In contrast, clinically established markers showed limited and inconsistent performance in the colorectal cohort. AMY2A showed only small fold changes from POD1 to POD3, with substantial overlap between leak and non-leak patients in the abundance distributions (Figure 4b). Its best early diagnostic performance was modest, with an AUC of 0.713 on POD1 and a wide confidence interval, falling below the performance reported in cohorts with established leakage.^33,34^ Although AMY2A increased on POD5, this late change should be interpreted cautiously because of reduced sample availability, particularly in the leak group. CRP showed low and inconsistent fold changes across postoperative days, with overlapping abundance distributions and no stable distinction between leak and non-leak patients (Figure 4d) consistent with variable performance reported.^35–37^ IL6 showed a slight increase at intermediate postoperative days, most visibly around POD3 to POD4 (Figure 4a), but patient-level distributions remained overlapping and did not indicate robust separation (Figure 4c). Contrary to the pancreatic cohort, PNLIP did not show a significant difference between two patient groups on any POD.

**Figure 4:**
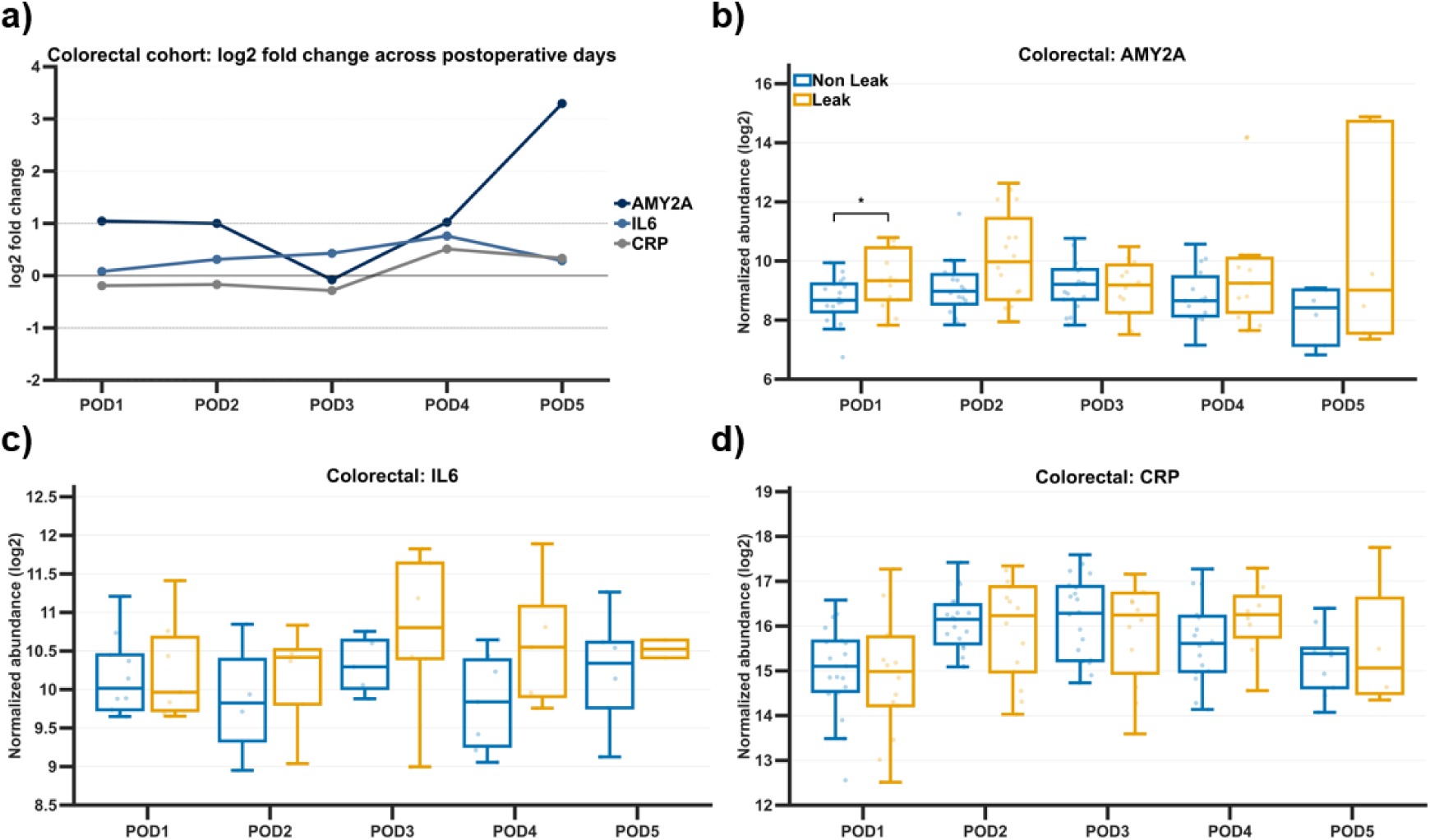
Temporal behavior and diagnostic performance established biomarkers in the colorectal cohort. a) Log2 fold change between leak and non-leak samples across postoperative days for AMY2A, CRP, and IL6. b–d) Distributions of normalized protein abundance for individual markers across postoperative days, shown separately for leak and non-leak samples. All markers show substantial overlap between groups across postoperative days. (* p<0.05)

Direct comparison of the two cohorts highlights a clear surgical-site-specific difference in the performance of the established biomarker panel evaluated here. In pancreatic surgery, AMY2A and PNLIP reflect pancreatic enzyme release and showed strong early discrimination between leak and non-leak patients with inflammatory biomarker increase in the following days. In colorectal surgery, no comparable dominant organ-specific signal was captured by the established markers included in this analysis. Instead, AMY2A, CRP, and IL6 showed overlapping distributions and inconsistent temporal behavior, indicating that this predefined marker panel has limited ability to distinguish colorectal leak from non-leak patients. Importantly, even in the pancreatic cohort, where established enzyme markers performed well, this panel provides only a limited view of leakage-associated proteomic changes. Additional discovery-based analysis is therefore required in both cohorts to identify whether further proteins or protein patterns contribute relevant information beyond the established biomarkers.

### Novel Biomarker Discovery

We next explored the full drain-fluid proteome to determine which additional leakage-associated signals could be identified beyond the clinically established biomarker panel. Discovery analysis was performed separately in both cohorts and was based on differential abundance screening using FDR ≤ 0.10 and |log2 fold change| ≥ 1, followed by prioritization according to recurrence across postoperative days, direction consistency, and effect size. This strategy was designed to emphasize longitudinally reproducible protein patterns rather than isolated single-day differences. Full ranked discovery lists are provided in Supplementary Tables S1 for the pancreatic cohort and S2 for the colorectal cohort.

In the pancreatic cohort, between 2781 and 3439 proteins were quantified per postoperative day, of which 466 met the differential expression criteria on at least one day across POD1 to POD8. Restricting the analysis to proteins with recurrent and directionally consistent differences across POD1 to POD5 yielded 277 discovery candidates, including 202 proteins enriched in leak samples and 54 enriched in non-leak samples. The temporal heatmap showed separation already at POD1, dominated by large positive fold changes of pancreatic enzymes^24,38^, including AMY2A, AMY2B, CPA1, PNLIP, PRSS1, and CPB1 (Figure 5a). These proteins formed a high-intensity leak-enriched band during the earliest postoperative phase, with effect sizes exceeding those of other protein groups.

**Figure 5:**
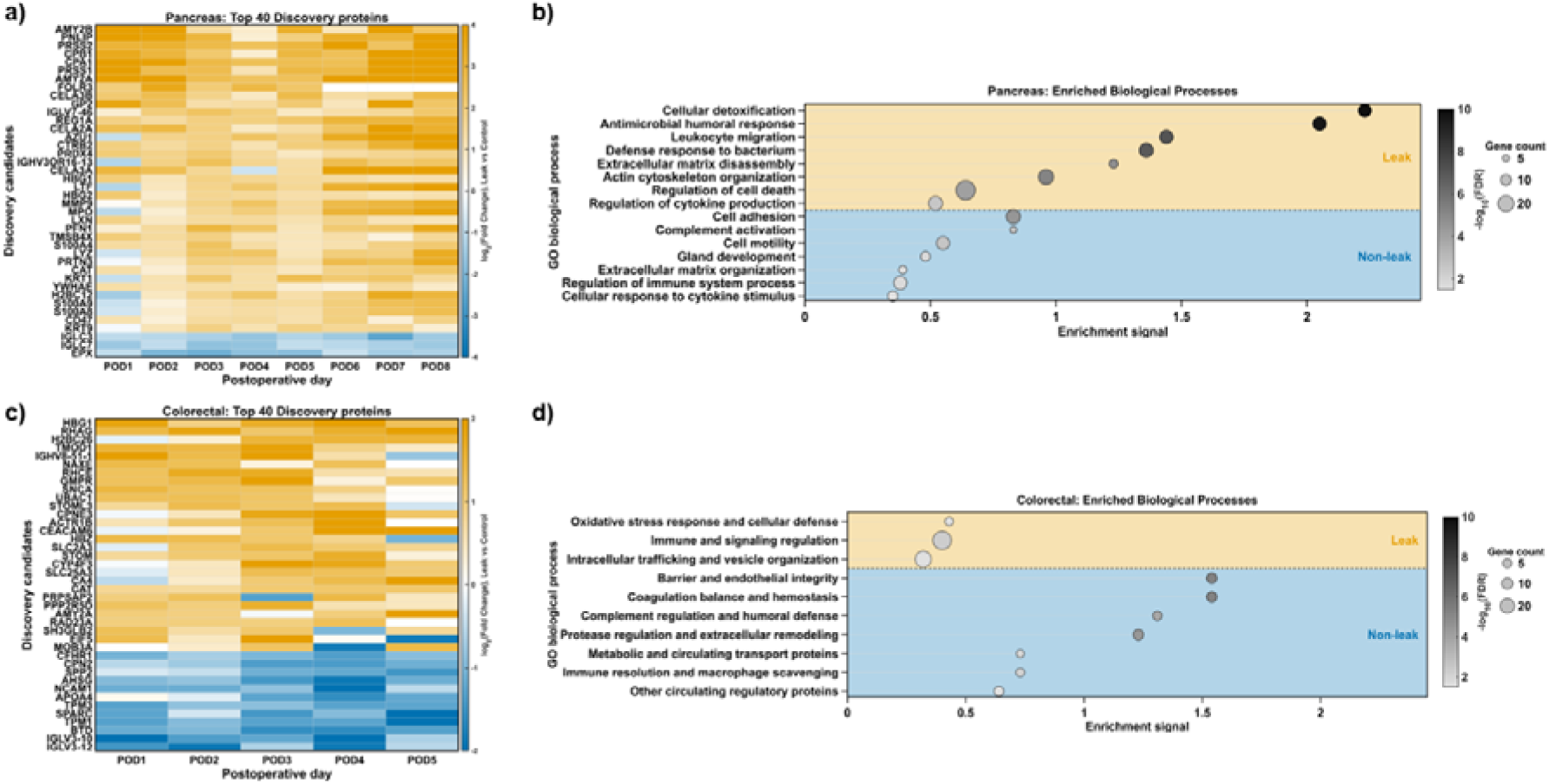
Temporal discovery patterns and functional enrichment in pancreatic and colorectal cohorts. a) Heatmap showing temporal differential expression of the top 40 discovery proteins in the pancreatic cohort across postoperative days (POD1–POD8). Values represent log2 fold change (leak vs non-leak). Orange indicates higher abundance in leaks, blue indicates higher abundance in non-leaks. Early strong positive fold changes at POD1 are dominated by pancreatic enzymes, followed by sustained elevation of additional proteins across POD2–POD4. (b) Gene ontology enrichment analysis of pancreatic discovery proteins. Each dot represents an enriched biological process, plotted by enrichment signal. Dot size indicates gene count and color represents −log10(FDR). Processes enriched in leak are shown in the upper panel and include antimicrobial response, leukocyte migration and extracellular matrix disassembly. Processes enriched in non-leaks are shown in the lower panel and include complement activation, cell adhesion and extracellular matrix organization. (c) Heatmap showing temporal differential expression of top 40 discovery proteins in the colorectal cohort across POD1–POD5. Compared to the pancreatic cohort, separation is limited at POD1–POD2 and becomes more pronounced at POD3, primarily driven by reduced abundance of proteins in leak patients. (d) Gene ontology enrichment analysis of colorectal discovery proteins. Non-leak enriched processes include barrier and endothelial integrity, extracellular matrix organization, complement regulation and coagulation related pathways. Leak enriched processes are less prominent and include oxidative stress response and intracellular trafficking. Dot size indicates gene count and color represents −log10(FDR).

Beyond this early enzyme-dominated pattern, additional proteins remained elevated in leak patients across POD2 to POD4 (median day of leak diagnosis: POD3). Proteins elevated during this phase were mainly associated with neutrophil activity, antimicrobial defense, oxidative stress regulation, and extracellular matrix remodelling^39,40^, with representative examples including AZU1, LYZ, S100A8/S100A9, MPO, and MMP9. By contrast, proteins enriched in non-leak patients formed a more stable pattern across postoperative days and were linked to complement activity, adhesion, and extracellular matrix organization. These patterns were supported by functional enrichment analysis, which separated leak-associated processes such as cellular detoxification, antimicrobial response, leukocyte migration, and extracellular matrix disassembly from non-leak-associated processes related to complement activation, cell adhesion, extracellular matrix organization, and immune regulation (Figure 5b).

The colorectal cohort showed a distinct discovery pattern. Instead of an early enzyme-dominated signal, the top-ranked colorectal candidates showed strongest group divergence around POD3 (Figure 5c), preceding the median clinical diagnosis of leakage in this cohort (median POD 5.5). This places the strongest proteomic separation within an early postoperative window before typical clinical detection. Of 2397 proteins included in the merged POD1–POD5 dataset, 270 met the differential expression criteria on at least one postoperative day. The longitudinal discovery analysis identified 214 candidates, including 163 proteins enriched in leak samples and 51 enriched in non-leak samples. Several of the most consistent non-leak-enriched candidates were associated with endothelial barrier integrity, extracellular matrix organization, complement regulation, coagulation balance, and immune regulation. Their lower abundance in leak patients suggests that colorectal leakage is characterized in part by reduced levels of proteins associated with uncomplicated recovery, rather than by the emergence of a single dominant leakage-derived marker. Leak-enriched colorectal candidates were comparatively fewer and less specific. Functional enrichment analysis supported this structure, showing a smaller leak-associated program related to oxidative stress, immune signaling, and vesicle organization, alongside a broader non-leak-associated faction consistent with tissue stability and postoperative recovery (Figure 5d).

Temporal analysis further distinguished the two surgical settings. In the pancreatic cohort, separation was already present at POD1 and remained evident across early postoperative days, whereas the colorectal cohort showed limited early separation and a distinct peak around POD3. This difference in timing suggests that the two proteomic profiles capture different stages of leakage-associated biology. In pancreatic surgery, sustained increases in protein abundance are consistent with ongoing exposure of the postoperative compartment to enzyme-rich pancreatic fluid^25,27,38^. In colorectal surgery, separation was driven in part by transient reduction of proteins associated with uncomplicated recovery, with maximal divergence around POD3, suggesting disruption or absence of stable healing before clinical manifestation^41–43^.

These findings also have implications for biomarker development. While biomarker studies often focus on proteins that increase during disease^10,16,24,44^, the colorectal cohort demonstrates that reduced abundance of recovery-associated proteins can provide strong separation between groups. Together, the discovery analyses argue against a universal biomarker strategy for postoperative leakage. In pancreatic surgery, candidate markers largely reflect the presence of leakage and associated tissue response. In colorectal surgery, an important component of the signal reflects loss of a stable recovery profile, indicating that monitoring recovery-associated proteins may be as informative as detecting leakage-associated increases. Biomarker panels could therefore be adapted to the biological context of each surgical procedure.

### Candidate Marker Selection for Future Diagnostics

Having established that discovery analysis identified procedure-specific protein patterns beyond the clinically validated biomarker panel, we next selected representative candidates for potential future validation in independent cohort studies. The aim was not to establish a final diagnostic panel at this stage, but to highlight proteins that captured the main biological and temporal features of each cohort and showed measurable separation between leak and non-leak patients over postoperative time.

For the pancreatic cohort, PRSS2 and AZU1 were selected to represent two complementary components of the leakage-associated profile. PRSS2 was chosen as a pancreatic digestive protease that relates directly to exocrine enzyme leakage, but has been far less frequently evaluated than established drain-fluid markers such as amylase or lipase, with only limited reports on trypsin-based AL detection in the literature.^45^ Although its discrimination was weaker than AMY2A or PNLIP, PRSS2 showed reproducible separation during the early postoperative phase, with AUC values of 0.78 on POD1 and 0.81 on POD2 (Figure 6a,c). AZU1 was selected as a representative discovery candidate from the broader inflammatory and neutrophil-associated response identified in the pancreatic analysis. In contrast to PRSS2, AZU1 showed increasing separation after POD1, with AUC values of 0.88 on POD2, 0.88 on POD3, and 0.91 on POD4 (Figure 6a,c). This temporal pattern suggests that PRSS2 and AZU1 capture different aspects of the pancreatic leak-associated proteome, an early enzyme-related signal and a later inflammatory response.

**Figure 6:**
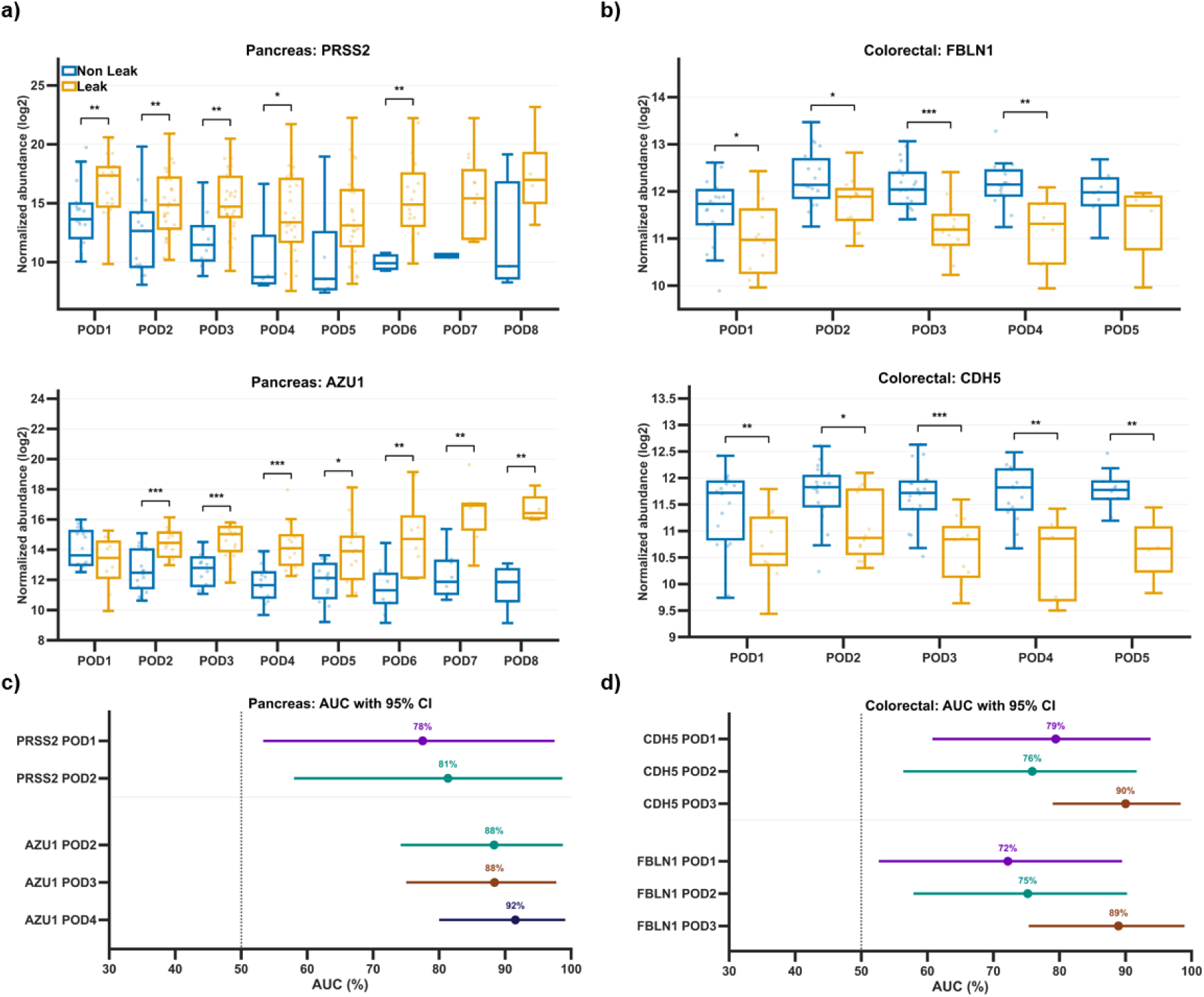
Temporal behavior of discovery biomarkers in the pancreatic and colorectal cohort. a) Log2 fold changes between leak and non-leak samples across postoperative days for PRSS2 and AZU1 in the pancreatic cohort. b) Log2 fold changes between leak and non-leak samples across postoperative days for FBLN1 and CDH5 in the colorectal cohort. c) Receiver operating characteristic curves for pancreatic discovery markers across significant PODs. d) Receiver operating characteristic curves for colorectal discovery markers across significant PODs.

For the colorectal cohort, FBLN1 and CDH5 were selected because they represented a prominent non-leak-enriched discovery pattern observed in this surgical setting. Both proteins are associated with tissue structure and vascular or extracellular matrix integrity, and both showed lower abundance in leak patients compared with non-leak patients across early postoperative days (Figure 6b). Their diagnostic performance was strongest around POD3, matching the time point of maximal colorectal proteomic divergence in the discovery analysis. CDH5 reached an AUC of 0.90 on POD3, while FBLN1 reached an AUC of 0.89 on POD3 (Figure 6d). These results support the interpretation that colorectal leakage-associated separation is driven by reduced abundance of proteins linked to uncomplicated recovery rather than by strong increases in leakage-derived proteins. Together, these four candidates were chosen to represent the procedure-specific structure revealed by the discovery analysis. In the pancreatic cohort, PRSS2 and AZU1 capture enzyme-associated and inflammatory components of leakage. In the colorectal cohort, FBLN1 and CDH5 capture the loss of recovery-associated protein patterns that distinguish leak from non-leak patients. Their temporal behavior and ROC performance support further targeted assessment in separate future prospective validation studies.

## CONCLUSION

This study demonstrates that longitudinal proteomic profiling of postoperative drain fluid captures distinct leakage-associated patterns in pancreatic and colorectal surgery. Although both complications are clinically classified as postoperative leaks, their proteomic signatures differed in timing, directionality, and biological composition. Pancreatic leakage was characterized by early enrichment of pancreatic enzymes together with inflammatory and tissue injury-associated proteins, whereas colorectal leakage showed a more temporally restricted divergence around POD3, driven largely by reduced abundance of proteins associated with uncomplicated postoperative recovery.

These findings show that established biomarker strategies may not be directly transferable across surgical settings. Enzyme-based markers such as AMY2A and PNLIP provided strong early discrimination in pancreatic surgery, while the predefined marker panel showed limited performance in the colorectal cohort. In contrast, discovery analysis identified procedure-specific candidate markers, including PRSS2 and AZU1 in pancreatic surgery and FBLN1 and CDH5 in colorectal surgery, which reflect different aspects of leakage-associated proteomic change and may provide a new outlook on the abdominal drain fluid proteome.

Overall, the results argue against a universal biomarker strategy for postoperative leakage and support procedure-specific approaches adapted to the underlying biology of each surgical context. Drain-fluid proteomics provides a local molecular readout of postoperative recovery and leakage development and may help identify early changes before routine clinical diagnosis. Future targeted validation in larger cohorts will be required to confirm the selected candidates, establish clinically useful concentration thresholds, and determine whether multi-protein panels improve early postoperative leak detection.

## METHODS

### Study Design and Cohorts

This study analyzed longitudinal drain fluid proteomes from two independent surgical cohorts representing distinct gastrointestinal procedures. The colorectal cohort was collected at the University Hospital in Pilsen, the study was conducted with approval of the ethics committee of the University Hospital in Pilsen (Ref. No.: 509/2020 and Ref. No.: 50/24). The pancreatic cohort was collected at the Cantonal Hospital St. Gallen (HOCH - Health Ostschweiz) with an ethics approval in place (BASEC no. Req-2022-01026 and BASEC no. Req-2026-01175). Patients were classified into leak and non-leak groups based on postoperative clinical outcome. Sample availability across postoperative days is summarized in Supplementary Table S3, with the colorectal cohort including samples from POD1 to POD5 and the pancreatic cohort including samples from POD1 to POD10. All samples were analyzed in anonymized form and linked to clinical annotations through coded identifiers.

### Leak Definitions and Clinical Classification

Leak status was assigned separately for each surgical cohort according to procedure-specific clinical criteria and documented postoperative outcome. The day of clinical diagnosis was defined as the first postoperative day on which leakage was documented by clinical assessment, drain-fluid biochemistry, imaging, or need for radiological or surgical intervention.

### Drain Fluid Collection and Sample Preparation for Mass Spectrometry–Based Proteomic Analysis

Postoperative drain fluid was collected from abdominal drains placed during surgery as part of routine postoperative management. Samples were assigned to PODs according to the day of collection following surgery.

Drain fluid samples were processed locally and stored frozen until analysis. Aliquots prepared for proteomic measurement were transferred to the Functional Genomics Center Zurich (FGCZ) for mass spectrometry analysis. Briefly, 40 µl drain-fluid was added to 10 µl pre-aliquoted 20 % sodium dodecyl sulfate (SDS) in 100 mM triethylammonium bicarbonate (TEAB), pH 8.2 to reach a final concentration of 4% SDS. For protein lysis, samples were incubated at 95 °C for 10 minutes, kept frozen for sample hand-over, and re-boiled for 10min before diluting to 2% SDS. Protein concentration was determined using the Lunatic UV/Vis polychromatic spectrophotometer (Unchained Labs) and based on an average concentration, 50 µg of proteins were taken for further processing. Proteins were reduced with 5 mM TCEP(tris(2-carboxyethyl)phosphine) and alkylated with 15 mM chloroacetamide at 30°C for 30 min. Samples were processed using the single pot solid phase enhanced sample preparation (SP3). The SP3 protein purification, digest and peptide clean-up were performed using a KingFisher Flex System (Thermo Fisher Scientific) and Carboxylate-Modified Magnetic Particles (GE Life Sciences; GE65152105050250, GE45152105050250).^46,47^ Beads were conditioned following the manufacturer’s instructions, consisting of 3 washes with water at a concentration of 1 µg/µl. Samples were diluted with 100% ethanol to a final concentration of 60% ethanol. The beads, wash solutions and samples were loaded into 96 deep well- or micro-plates and transferred to the KingFisher. Following steps were carried out on the robot: collection of beads from the last wash, protein binding to beads, washing of beads in wash solutions 1-3 (80% ethanol), protein digestion (overnight at 37°C with a trypsin:protein ratio of 1:50 in 50 mM Triethylammoniumbicarbonat (TEAB)) and peptide elution from the magnetic beads using MilliQ water. The digest solution and water elution were combined and acidified. For each sample an amount according to 2 absorbance (Lunatic, unmix) were loaded onto Evotips (Evosep Biosystems), according to the manufacturer’s instructions.

### LC-MSMS – timsTOF Evosep diaPASEF

MS analyses were performed on a timsTOF Pro (colorectal cohort) and timsTOF Flex (pancreatic cohort) (Bruker) coupled to an Evosep One (EvoSep Biosystems). Samples were separated with the extended Evosep method ‘’30 samples/day’’ keeping the analytical column (PepSep C18, 15 cm x 150 µm, 1.5 µm) at 50°C. For the dual timsTOF, MS spectra were scanned from m/z 100 to m/z 1700 and inverse mobilities [1/K0] from 0.60 Vs/cm2 to 1.60 Vs/cm2 with an ion accumulation and ramp time of 100 ms, respectively. The data were acquired in diaPASEF mode (data independent acquisition Parallel Accumulation Serial Fragmentation); 1 MS scan was followed by 16 PASEF cycles between m/z 400 to m/z 1200 with overlapping isolation windows of m/z 26, covering with 2 x 0.30 [1/K0] windows the ion mobility range from 0.60 Vs/cm2 to 1.42 Vs/cm2. Singly charged ions were excluded using the polygon filter mask.

The mass spectrometry proteomics data was handled using the local laboratory information management system (LIMS)^48^ and all relevant data have been deposited to the ProteomeXchange Consortium via the PRIDE (http://www.ebi.ac.uk/pride) partner repository with the data set identifier PXD082170 and PXD082236.

### Protein Identification and Quantification

Raw mass spectrometry data were processed using DIA-NN version 1.8.2 (colorectal) and 1.9.2 (pancreatic) in library-free mode.^49^ Searches were performed against the UniProt human reference proteome database (UP000005640, one protein sequence per gene) supplemented with common contaminants, using precursor charges +2 and +3, precursor mass range 400-1500 m/z, fragment mass range 200-1800 m/z, MS1 and MS2 mass accuracies of 15 ppm and 20 ppm, respectively, trypsin/P specificity with one missed cleavage allowed, and methionine oxidation as variable modification. The maximum false discovery rate was set to 0.01. DIA-NN report files containing precursor ion abundances for each raw file were used as input for downstream protein-level quantification and statistical analysis.

### Differential Protein Analysis

Differential protein abundance between leak and non-leak patients was evaluated separately for each POD using the prolfqua R package.^50^ Starting from DIA-NN precursor-level report files, precursor abundances were first aggregated to peptidoform abundances and then to protein abundances using Tukey’s median polish. Protein abundances were transformed using variance-stabilizing normalization before fitting linear models.^51^ Statistical testing was used to estimate group differences, confidence intervals, and false discovery rates for all quantifiable proteins; proteins were classified as differentially expressed when two criteria were met: FDR <= 0.10 and absolute log2 fold change >= 1. This approach enabled identification of proteins consistently enriched in leak or non-leak samples across PODs. For longitudinal candidate selection, proteins were required to meet the differential-expression criteria on at least one postoperative day and to show a consistent direction of regulation, defined by the same sign of the log2 fold change, on at least four of POD1–POD5. Candidates were subsequently prioritized according to the number of significant PODs, consistency of the log2 fold-change direction, median absolute effect size, and best per-POD FDR (lowest).

### Functional Enrichment and Interaction Analysis

Functional enrichment and protein interaction analyses were performed using the STRING database.^52^ Lists of differentially expressed proteins were analyzed for enrichment of Gene Ontology (GO) biological processes and for protein–protein interaction networks. Network connectivity was assessed by comparing the observed number of protein–protein interaction edges with the number expected by chance. This analysis was used to determine whether differentially expressed proteins formed coordinated biological pathways rather than isolated molecular signals.

### Biomarker Evaluation using ROC Analysis

Receiver operating characteristic (ROC) analysis was performed to evaluate the diagnostic discrimination between leak and non-leak samples for selected proteins.^53^ To ensure statistical stability, ROC analyses were restricted to PODs with at least ten patients per group.

For each eligible protein and POD, the area under the ROC curve (AUC) and corresponding confidence intervals (CI) were calculated. These metrics were used to evaluate the diagnostic performance of candidate biomarkers.

## Data Availability

The full proteomic data is available on the PRIDE database PXD082170 and PXD082236. Any additional information and data is available upon reasonable request to the corresponding author.

https://www.ebi.ac.uk/pride/archive/projects/PXD082170/public

https://www.ebi.ac.uk/pride/archive/projects/PXD082236/public

## AUTHOR CONTRIBUTIONS

I.K.H. conceived the project idea. A.J. planned and coordinated the experiments, helped with data evaluation and wrote the manuscript. V.M.A. evaluated the proteomic data and gave feedback on the manuscript. S.P. performed the measurements at the FGCZ, helped with data analysis and gave feedback on the manuscript. I.T. planned and conducted the sample collection of the pancreatic cohort, gave clinical insight and feedback on the manuscript. V.L. and R.P. planned and conducted the sample collection of the colorectal cohort, gave clinical insight and feedback on the manuscript. All authors reviewed, contributed, and approved the final version of the manuscript.

## ACKNOWLEDGEMENTS

We kindly thank Witold Wolski for providing answers to our data analysis questions and his work in preparing the data at the FGCZ and for providing feedback on the manuscript. We kindly acknowledge funding from the Swiss National Science Foundation (Eccellenza grant no. 181290 and Project grant no. IC00I0L-227785 to I.K.H.), the Evi Diethelm-Winteler-Stiftung (I.K.H.) the ETH Domain Specific Focus Area Personalized Health and Related Technologies (PHRT) program (grant number 2023/982 to I.K.H.) and the Swiss National Science Foundation BRIDGE Discovery Program (grant number 40B2-0_226647 to I.K.H.).

## List of Abbreviations

General abbreviations

AL: Anastomotic leakage
AUC: Area under the curve
CI: Confidence interval
FDR: False discovery rate
FGCZ: Functional Genomic Center Zurich
GO: Gene Ontology
POD: Postoperative day
ROC: Receiver operating characteristic

Protein nomenclature

Protein: Full Name
AMY2A: Amylase alpha 2A
C1QA: Complement component 1q subcomponent A chain
C1QB: Complement component 1q subcomponent B chain
C1S: Complement component 1s
CDH5: Cadherin 5, vascular endothelial cadherin
CFH: Complement factor H
CFHR1: Complement factor H related protein 1
CPNE3: Copine 3
CRP: C-reactive protein
DAG1: Dystroglycan 1
FBLN1: Fibulin 1
IL6: Interleukin 6
LAMC1: Laminin subunit gamma 1
LYZ: Lysozyme
MMP8: Matrix metalloproteinase 8
MPO: Myeloperoxidase
PARK7: Parkinson protein 7, DJ-1
PI16: Peptidase inhibitor 16
PNLIP: Pancreatic lipase
PRSS2: Protease serine 2, trypsin 2
PROS1: Protein S
RAB27A: Ras related protein Rab 27A
RAB5B: Ras related protein Rab 5B
S100A8: S100 calcium binding protein A8
S100A9: S100 calcium binding protein A9
TNXB: Tenascin XB
VCAM1: Vascular cell adhesion molecule 1

## References

(1) Bashir Mohamed, K.; Hansen, C. H.; Krarup, P.-M.; Fransgård, T.; Madsen, M. T.; Gögenur, I. The Impact of Anastomotic Leakage on Recurrence and Long-Term Survival in Patients with Colonic Cancer: A Systematic Review and Meta-Analysis. European Journal of Surgical Oncology 2020, 46 (3), 439–447. 10.1016/j.ejso.2019.10.038

(2) Turrentine, F. E.; Denlinger, C. E.; Simpson, V. B.; Garwood, R. A.; Guerlain, S.; Agrawal, A.; Friel, C. M.; LaPar, D. J.; Stukenborg, G. J.; Jones, R. S. Morbidity, Mortality, Cost, and Survival Estimates of Gastrointestinal Anastomotic Leaks. Journal of the American College of Surgeons 2015, 220 (2), 195–206. 10.1016/j.jamcollsurg.2014.11.002

(3) Ju, J.-W.; Jang, H. S.; Lee, M.; Lee, H.-J.; Kwon, W.; Jang, J.-Y. Early Postoperative Fever as a Predictor of Pancreatic Fistula after Pancreaticoduodenectomy: A Single-Center Retrospective Observational Study. BMC Surg 2024, 24 (1), 229. 10.1186/s12893-024-02521-0

(4) Gielen, A. H. C.; Heuvelings, D. J. I.; Sylla, P.; van Loon, Y.-T.; Melenhorst, J.; Bouvy, N. D.; Kimman, M. L.; Breukink, S. O.; Collaborative, O. behalf of the C. Impact of Anastomotic Leakage After Colorectal Cancer Surgery on Quality of Life: A Systematic Review. Diseases of the Colon & Rectum 2025, 68 (2), 154. 10.1097/DCR.0000000000003478

(5) Leourier, P.; Pellegrin, A.; Regimbeau, J.-M.; Sabbagh, C. Is Early CT in Cases of Elevated Postoperative CRP the Best Option for the Diagnosis of Colorectal Anastomotic Leakage? Int J Colorectal Dis 2023, 38 (1), 278. 10.1007/s00384-023-04571-x

(6) Sutcliffe, R. P.; Battula, N.; Haque, A.; Ali, A.; Srinivasan, P.; Atkinson, S. W.; Rela, M.; Heaton, N. D.; Prachalias, A. A. Utility of Drain Fluid Amylase Measurement on the First Postoperative Day after Pancreaticoduodenectomy. World Journal of Surgery 2012, 36 (4), 1. 10.1007/s00268-012-1460-0

(7) Bassi, C.; Marchegiani, G.; Dervenis, C.; Sarr, M.; Abu Hilal, M.; Adham, M.; Allen, P.; Andersson, R.; Asbun, H. J.; Besselink, M. G.; Conlon, K.; Del Chiaro, M.; Falconi, M.; Fernandez-Cruz, L.; Fernandez-del Castillo, C.; Fingerhut, A.; Friess, H.; Gouma, D. J.; Hackert, T.; Izbicki, J.; Lillemoe, K. D.; Neoptolemos, J. P.; Olah, A.; Schulick, R.; Shrikhande, S. V.; Takada, T.; Takaori, K.; Traverso, W.; Vollmer, C.; Wolfgang, C. L.; Yeo, C. J.; Salvia, R.; Buchler, M. The 2016 Update of the International Study Group (ISGPS) Definition and Grading of Postoperative Pancreatic Fistula: 11 Years After. Surgery 2017, 161 (3), 584–591. 10.1016/j.surg.2016.11.014

(8) Bertens, K. A.; Crown, A.; Clanton, J.; Alemi, F.; Alseidi, A. A.; Biehl, T.; Helton, W. S.; Rocha, F. G. What Is a Better Predictor of Clinically Relevant Postoperative Pancreatic Fistula (CR-POPF) Following Pancreaticoduodenectomy (PD): Postoperative Day One Drain Amylase (POD1DA) or the Fistula Risk Score (FRS)? HPB 2017, 19 (1), 75–81. 10.1016/j.hpb.2016.10.001

(9) Villafane-Ferriol, N.; Van Buren, G.; Mendez-Reyes, J. E.; McElhany, A. L.; Massarweh, N. N.; Silberfein, E. J.; Hsu, C.; Tran Cao, H. S.; Schmidt, C.; Zyromski, N. J.; Dillhoff, M. E.; Roch, A.; Oliva, E.; Smith, A. C.; Zhang, Q.; Fisher, W. E. Sequential Drain Amylase to Guide Drain Removal Following Pancreatectomy. HPB 2018, 20 (6), 514–520. 10.1016/j.hpb.2017.11.008

(10) Clark, D. A.; Steffens, D.; Solomon, M. An Umbrella Systematic Review of Drain Fluid Analysis in Colorectal Surgery for the Detection of Anastomotic Leak: Not yet Ready to Translate Research Studies into Clinical Practice. Colorectal Disease 2021, 23 (11), 2795–2805. 10.1111/codi.15844

(11) Clark, D. A.; Edmundson, A.; Steffens, D.; Harris, C.; Stevenson, A.; Solomon, M. Drain Fluid Amylase as a Biomarker for the Detection of Anastomotic Leakage after Rectal Resection without a Diverting Ileostomy. ANZ Journal of Surgery 2022, 92 (4), 813–818. 10.1111/ans.17461

(12) Matthiessen, P.; Strand, I.; Jansson, K.; Törnquist, C.; Andersson, M.; Rutegård, J.; Norgren, L. Is Early Detection of Anastomotic Leakage Possible by Intraperitoneal Microdialysis and Intraperitoneal Cytokines after Anterior Resection of the Rectum for Cancer? Dis Colon Rectum 2007, 50 (11), 1918–1927. 10.1007/s10350-007-9023-4

(13) El Zaher, H. A.; Ghareeb, W. M.; Fouad, A. M.; Madbouly, K.; Fathy, H.; Vedin, T.; Edelhamre, M.; Emile, S. H.; Faisal, M. Role of the Triad of Procalcitonin, C-Reactive Protein, and White Blood Cell Count in the Prediction of Anastomotic Leak Following Colorectal Resections. World J Surg Onc 2022, 20 (1), 33. 10.1186/s12957-022-02506-4

(14) Klupp, F.; Schuler, S.; Kahlert, C.; Halama, N.; Franz, C.; Mayer, P.; Schmidt, T.; Ulrich, A. Evaluation of the Inflammatory Markers CCL8, CXCL5, and LIF in Patients with Anastomotic Leakage after Colorectal Cancer Surgery. Int J Colorectal Dis 2020, 35 (7), 1221–1230. 10.1007/s00384-020-03582-2

(15) Kyrochristou, I.; Anagnostopoulos, G.; Psalla, K.; Giannakakis, P.; Rogdakis, A. Diagnostic Accuracy of CRP in the Drainage Fluid for Early Detection of Anastomotic Leakage in Colorectal Surgery – a Pilot Study. Folia Medica 2025, 67 (4), e154087. 10.3897/folmed.67.e154087

(16) Su’a, B. U.; Mikaere, H. L.; Rahiri, J. L.; Bissett, I. B.; Hill, A. G. Systematic Review of the Role of Biomarkers in Diagnosing Anastomotic Leakage Following Colorectal Surgery. Br J Surg 2017, 104 (5), 503–512. 10.1002/bjs.10487

(17) Fukada, M.; Murase, K.; Higashi, T.; Yasufuku, I.; Sato, Y.; Tajima, J. Y.; Kiyama, S.; Tanaka, Y.; Okumura, N.; Takahashi, T.; Matsuhashi, N. Drain Fluid and Serum Amylase Concentration Ratio Is the Most Reliable Indicator for Predicting Postoperative Pancreatic Fistula after Distal Pancreatectomy. BMC Surg 2023, 23, 87. 10.1186/s12893-023-01980-1

(18) Schots, J. P. M.; Luyer, M. D. P.; Nieuwenhuijzen, G. A. P. Abdominal Drainage and Amylase Measurement for Detection of Leakage After Gastrectomy for Gastric Cancer. J Gastrointest Surg 2018, 22 (7), 1163–1170. 10.1007/s11605-018-3789-7

(19) Lee, C. W.; Pitt, H. A.; Riall, T. S.; Ronnekleiv-Kelly, S. S.; Israel, J. S.; Leverson, G. E.; Parmar, A. D.; Kilbane, E. M.; Hall, B. L.; Weber, S. M. Low Drain Fluid Amylase Predicts Absence of Pancreatic Fistula Following Pancreatectomy. J Gastrointest Surg 2014, 18 (11), 1902–1910. 10.1007/s11605-014-2601-6

(20) Koek, S.; Wiegele, S.; Ballal, M. Drain Fluid Amylase and Lipase as a Predictive Factor of Postoperative Pancreatic Fistula. ANZ Journal of Surgery 2022, 92 (3), 414–418. 10.1111/ans.17296

(21) Facy, O.; Chalumeau, C.; Poussier, M.; Binquet, C.; Rat, P.; Ortega-Deballon, P. Diagnosis of Postoperative Pancreatic Fistula. Br J Surg 2012, 99 (8), 1072–1075. 10.1002/bjs.8774

(22) Shi, J.; Wu, Z.; Wu, X.; Shan, F.; Zhang, Y.; Ying, X.; Li, Z.; Ji, J. Early Diagnosis of Anastomotic Leakage after Colorectal Cancer Surgery Using an Inflammatory Factors-Based Score System. BJS Open 2022, 6 (3), zrac069. 10.1093/bjsopen/zrac069

(23) Qiao, X.; Qi, X.; Xing, P.; Liu, T.; Hao, H.; Yang, X.; Jiang, B.; Cui, M.; Su, X. Tandem Mass Tag-Based Proteomic Profiling Identifies Biomarkers in Drainage Fluid for Early Detection of Anastomotic Leakage after Rectal Cancer Resection. J. Proteome Res. 2023, 22 (11), 3559–3569. 10.1021/acs.jproteome.3c00394

(24) Wiedemann, D.; Strotmann, J.; Fahlbusch, T.; Majchrzak-Stiller, B.; Peters, I.; Uhl, W.; Höhn, P. Biochemical Early Detection of Postoperative Pancreatic Fistula. Visc Med 2025, 1–9. 10.1159/000545091

(25) Mintziras, I.; Maurer, E.; Kanngiesser, V.; Bartsch, D. K. C-Reactive Protein and Drain Amylase Accurately Predict Clinically Relevant Pancreatic Fistula after Partial Pancreaticoduodenectomy. International Journal of Surgery 2020, 76, 53–58. 10.1016/j.ijsu.2020.02.025

(26) Caputo, D.; Angeletti, S.; Ciccozzi, M.; Cartillone, M.; Cascone, C.; La Vaccara, V.; Coppola, A.; Coppola, R. Role of Drain Amylase Levels Assay and Routinary Postoperative Day 3 Abdominal CT Scan in Prevention of Complications and Management of Surgical Drains after Pancreaticoduodenectomy. Updates Surg 2020, 72 (3), 727–741. 10.1007/s13304-020-00784-9

(27) Koek, S.; Wiegele, S.; Ballal, M. Drain Fluid Amylase and Lipase as a Predictive Factor of Postoperative Pancreatic Fistula. ANZ Journal of Surgery 2022, 92 (3), 414–418. 10.1111/ans.17296

(28) Doussot, B.; Doussot, A.; Ayav, A.; Santucci, N.; Deguelte, S.; Sow, A. K.; El Amrani, M.; Duvillard, L.; Piessen, G.; Girard, E.; Mabrut, J.-Y.; Garnier, J.; Ortega-Deballon, P.; Fournel, I.; Facy, O. Diagnostic Accuracy of Lipase as Early Predictor of Postoperative Pancreatic Fistula: Results from the LIPADRAIN Study. Ann Surg Open 2024, 5 (3), e492. 10.1097/AS9.0000000000000492

(29) Chui, J. N.; Ziaziaris, W. A.; Nahm, C. B.; Fuchs, T.; Sahni, S.; Lim, C. S. H.; Gill, A. J.; Samra, J. S.; Mittal, A. Lipase-to-Amylase Ratio for the Prediction of Clinically Relevant Postoperative Pancreatic Fistula Following Pancreaticoduodenectomy. Pancreas 2024, 53 (7), e579. 10.1097/MPA.0000000000002345

(30) Tzedakis, S.; Sauvanet, A.; Schiavone, R.; Razafinimanana, M.; Cauchy, F.; Rouet, J.; Dousset, B.; Gaujoux, S. What Should We Trust to Define, Predict and Assess Pancreatic Fistula after Pancreatectomy? Pancreatology 2020, 20 (8), 1779–1785. 10.1016/j.pan.2020.10.036

(31) Malya, F. U.; Hasbahceci, M.; Tasci, Y.; Kadioglu, H.; Guzel, M.; Karatepe, O.; Dolay, K. The Role of C-Reactive Protein in the Early Prediction of Serious Pancreatic Fistula Development after Pancreaticoduodenectomy. Gastroenterology Research and Practice 2018, 2018 (1), 9157806. 10.1155/2018/9157806

(32) Gasteiger, S.; Primavesi, F.; Göbel, G.; Braunwarth, E.; Cardini, B.; Maglione, M.; Sopper, S.; Öfner, D.; Stättner, S. Early Post-Operative Pancreatitis and Systemic Inflammatory Response Assessed by Serum Lipase and IL-6 Predict Pancreatic Fistula. World Journal of Surgery 2020, 44 (12), 1. 10.1007/s00268-020-05768-9

(33) Amroun, K.; Deguelte, S.; Djerada, Z.; Ramont, L.; Perrenot, C.; Rached, L.; Renard, Y.; Rhaiem, R.; Kianmanesh, R. High Amylase Concentration in Drainage Liquid Can Early Predict Proximal and Distal Intestinal Anastomotic Leakages: A Prospective Observational Study. J Res Med Sci 2023, 28, 5. 10.4103/jrms.jrms_273_21

(34) Ishiyama, Y.; Hirano, Y.; Yamato, M.; Akuta, S.; Yoshizawa, M.; Fujii, T.; Okazaki, N.; Hiranuma, C.; Sakuramoto, S. Drainage Fluid Amylase as a Biomarker for the Detection of Anastomotic Leakage After Low Anterior Resection of Rectal Cancer: A Two-Center Study. Cancer Diagn Progn 2024, 4 (6), 802–807. 10.21873/cdp.10399

(35) Ortega-Deballon, P.; Radais, F.; Facy, O.; D’Athis, P.; Masson, D.; Charles, P. E.; Cheynel, N.; Favre, J.-P.; Rat, P. C-Reactive Protein Is an Early Predictor of Septic Complications after Elective Colorectal Surgery. World J Surg 2010, 34 (4), 808–814. 10.1007/s00268-009-0367-x

(36) Vun, T.; Wu, Z.; Chea, C.; Liu, W.; Tao, R.; Deng, Y. C-Reactive Protein in Peritoneal Fluid for Predicting Anastomotic Leakage After Colorectal Cancer Surgery: A Systematic Review and Meta-Analysis. J Clin Med 2025, 14 (6), 2099. 10.3390/jcm14062099

(37) Dost, W.; Rasully, M. Q.; Zaman, M. N.; Dost, W.; Ali, W.; Ayobi, S. A.; Dost, R.; Niazi, J.; Bakht, K.; Iqbal, A.; Bokhari, S. F. H. Predictive Biomarkers for the Early Detection of Anastomotic Leaks in Colorectal Surgeries: A Systematic Review. Cureus 2024. 10.7759/cureus.74616

(38) Wüster, C.; Shi, H.; Kühlbrey, C. M.; Biesel, E. A.; Hopt, U. T.; Fichtner-Feigl, S.; Wittel, U. A. Pancreatic Inflammation and Proenzyme Activation Are Associated With Clinically Relevant Postoperative Pancreatic Fistulas After Pancreas Resection. Annals of Surgery 2020, 272 (5), 863. 10.1097/SLA.0000000000004257

(39) Merza, M.; Hartman, H.; Rahman, M.; Hwaiz, R.; Zhang, E.; Renström, E.; Luo, L.; Mörgelin, M.; Regner, S.; Thorlacius, H. Neutrophil Extracellular Traps Induce Trypsin Activation, Inflammation, and Tissue Damage in Mice With Severe Acute Pancreatitis. Gastroenterology 2015, 149 (7), 1920–1931.e8. 10.1053/j.gastro.2015.08.026

(40) Ishqi, H. M.; Ali, M.; Dawra, R. Recent Advances in the Role of Neutrophils and Neutrophil Extracellular Traps in Acute Pancreatitis. Clin Exp Med 2023, 23 (8), 4107–4122. 10.1007/s10238-023-01180-4

(41) Thompson, S. K.; Chang, E. Y.; Jobe, B. A. Clinical Review: Healing in Gastrointestinal Anastomoses, Part I. Microsurgery 2006, 26 (3), 131–136. 10.1002/micr.20197

(42) Morgan, R. B.; Shogan, B. D. The Science of Anastomotic Healing. Semin Colon Rectal Surg 2022, 33 (2), 100879. 10.1016/j.scrs.2022.100879

(43) Guo, S.; DiPietro, L. A. Factors Affecting Wound Healing. J Dent Res 2010, 89 (3), 219–229. 10.1177/0022034509359125

(44) Gray, M.; Marland, J. R. K.; Murray, A. F.; Argyle, D. J.; Potter, M. A. Predictive and Diagnostic Biomarkers of Anastomotic Leakage: A Precision Medicine Approach for Colorectal Cancer Patients. J Pers Med 2021, 11 (6), 471. 10.3390/jpm11060471

(45) Watanabe, G.; Ishizawa, T.; Kuriki, Y.; Kamiya, M.; Ichida, A.; Kawaguchi, Y.; Akamatsu, N.; Kaneko, J.; Arita, J.; Kokudo, N.; Urano, Y.; Hasegawa, K. Evaluation of Pancreatic Chymotrypsin Activity for On-Site Prediction of Clinically Relevant Postoperative Pancreatic Fistula. Pancreatology 2024, 24 (1), 169–177. 10.1016/j.pan.2023.11.017

(46) Hughes, C. S.; Foehr, S.; Garfield, D. A.; Furlong, E. E.; Steinmetz, L. M.; Krijgsveld, J. Ultrasensitive Proteome Analysis Using Paramagnetic Bead Technology. Mol Syst Biol 2014, 10 (10), MSB145625. 10.15252/msb.20145625

(47) Leutert, M.; Rodríguez-Mias, R. A.; Fukuda, N. K.; Villén, J. R2-P2 Rapid-Robotic Phosphoproteomics Enables Multidimensional Cell Signaling Studies. Mol Syst Biol 2019, 15 (12), e9021. 10.15252/msb.20199021

(48) Türker, C.; Akal, F.; Joho, D.; Panse, C.; Barkow-Oesterreicher, S.; Rehrauer, H.; Schlapbach, R. B-Fabric: The Swiss Army Knife for Life Sciences. In Proceedings of the 13th International Conference on Extending Database Technology; ACM: Lausanne Switzerland, 2010; pp 717–720. 10.1145/1739041.1739135

(49) Demichev, V.; Messner, C. B.; Vernardis, S. I.; Lilley, K. S.; Ralser, M. DIA-NN: Neural Networks and Interference Correction Enable Deep Proteome Coverage in High Throughput. Nat Methods 2020, 17 (1), 41–44. 10.1038/s41592-019-0638-x

(50) Wolski, W. E.; Nanni, P.; Grossmann, J.; d’Errico, M.; Schlapbach, R.; Panse, C. Prolfqua: A Comprehensive R-Package for Proteomics Differential Expression Analysis. J. Proteome Res. 2023, 22 (4), 1092–1104. 10.1021/acs.jproteome.2c00441

(51) Huber, W.; von Heydebreck, A.; Sültmann, H.; Poustka, A.; Vingron, M. Variance Stabilization Applied to Microarray Data Calibration and to the Quantification of Differential Expression. Bioinformatics 2002, 18 (suppl_1), S96–S104. 10.1093/bioinformatics/18.suppl_1.S96

(52) Szklarczyk, D.; Kirsch, R.; Koutrouli, M.; Nastou, K.; Mehryary, F.; Hachilif, R.; Gable, A. L.; Fang, T.; Doncheva, N. T.; Pyysalo, S.; Bork, P.; Jensen, L. J.; von Mering, C. The STRING Database in 2023: Protein-Protein Association Networks and Functional Enrichment Analyses for Any Sequenced Genome of Interest. Nucleic Acids Res 2023, 51 (D1), D638–D646. 10.1093/nar/gkac1000

(53) Hanley, J. A.; McNeil, B. J. The Meaning and Use of the Area under a Receiver Operating Characteristic (ROC) Curve. Radiology 1982, 143 (1), 29–36. 10.1148/radiology.143.1.7063747

