## Supplementary Files for "Longitudinal Drain-Fluid Proteomics Reveals Procedure-Specific Signatures of Postoperative Leakage after Pancreatic and Colorectal Surgery"

**Supplementary Figure S1: Differential protein abundance across postoperative days in the pancreatic cohort.** Volcano plots showing log2 fold change between leak and non-leak samples and corresponding significance for each postoperative day. Each point represents one protein. Positive values indicate higher abundance in leak; negative values indicate higher abundance in non-leak. Vertical lines mark ±1 fold change and the horizontal line marks FDR ≤ 0.10. Proteins statistically higher in the leak group leak are shown in red and proteins higher in the non-leak group in green.

.

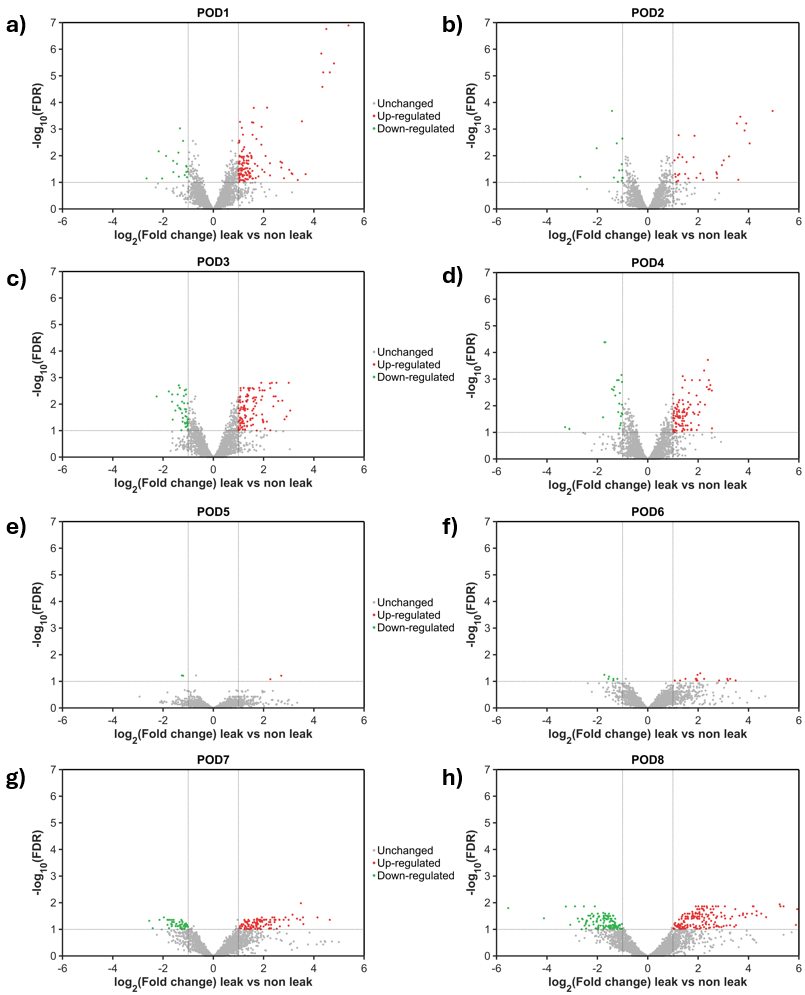

**Supplementary Figure S2: Differential protein abundance across postoperative days in the colorectal cohort.** Volcano plots showing log2 fold change between leak and non-leak samples and corresponding significance for each postoperative day. Each point represents one protein. Positive values indicate higher abundance in leak; negative values indicate higher abundance in non-leak. Vertical lines mark ±1 fold change and the horizontal line marks FDR ≤ 0.10. Proteins statistically higher in the leak group leak are shown in red and proteins higher in the non-leak group in green.

.

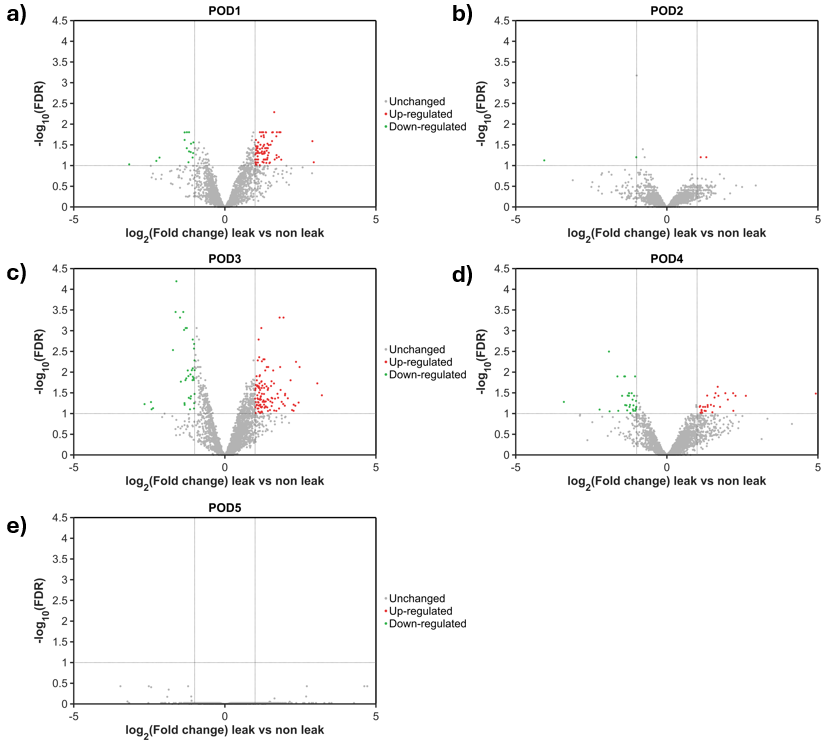

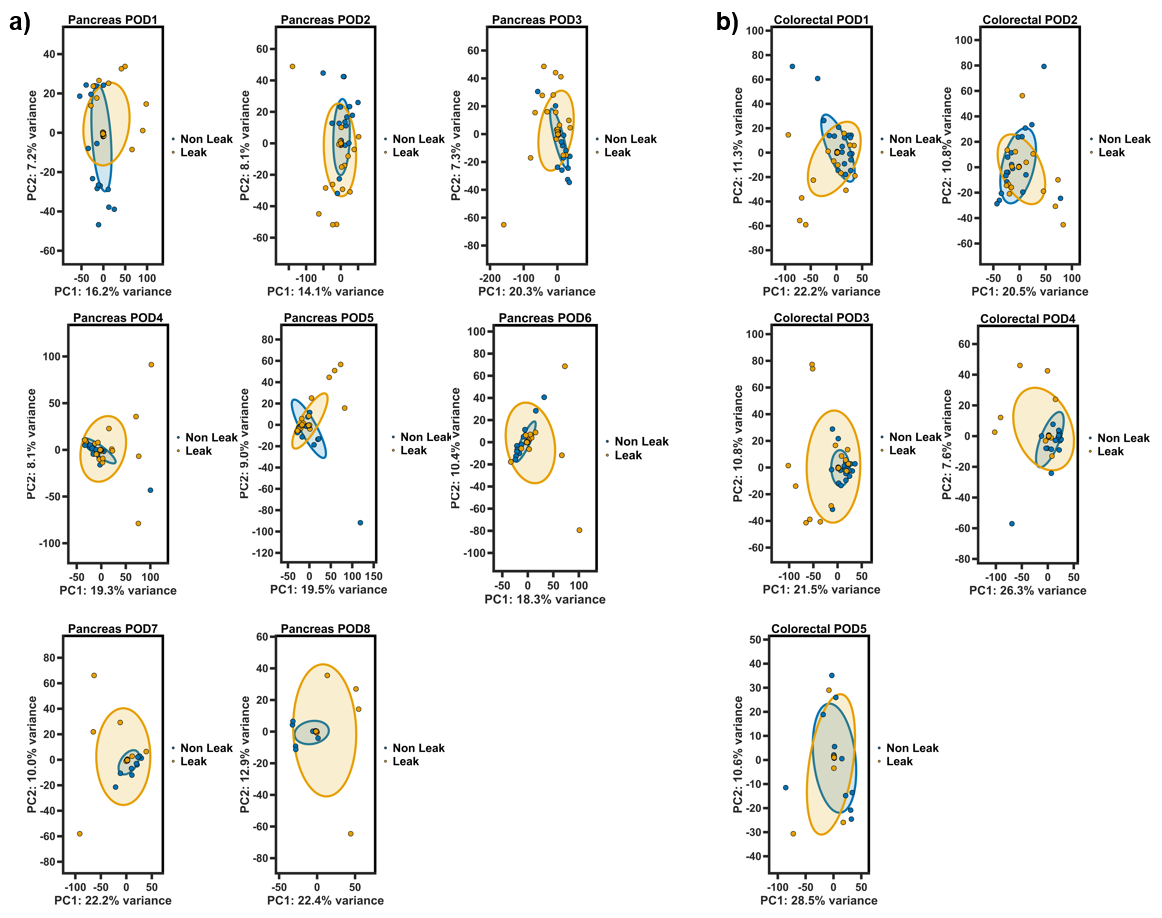

**Supplementary Figure S3: PCA analysis across postoperative days in both cohorts.** PCA plots show the distribution of leak and non-leak samples at individual postoperative days in the pancreatic cohort in a) (POD1–POD8) and colorectal cohort (POD1–POD5) in c). Each point represents an individual patient sample, with blue indicating non-leak and orange indicating leak patients. Ellipses summarize the distribution of each group. Axis labels indicate the proportion of total variance explained by the first two principal components.

**Supplementary Table S1: Full ranked list of differentially expressed proteins in the pancreatic cohort across postoperative days.**

| **Rank** | **Protein** | **BestPerPOD_FDR** | **MedLog2FC** | **PeakLog2FC** | **Sign. POD’s** | **Same Sign POD’s** | **Direction** |
| --- | --- | --- | --- | --- | --- | --- | --- |
| **1** | IGLC3 | 4.13E-05 | -1.25 | -1.68 | 5 | 5 | Non-Leak-Enriched |
| **2** | IGHG2 | 1.10E-03 | -1.22 | -1.63 | 5 | 5 | Non-Leak-Enriched |
| **3** | PRDX4 | 1.57E-04 | 1.86 | 2.25 | 4 | 5 | Leak-Enriched |
| **4** | IGLC7 | 4.13E-05 | -1.60 | -1.77 | 4 | 5 | Non-Leak-Enriched |
| **5** | S100A4 | 1.57E-03 | 1.46 | 2.51 | 4 | 5 | Leak-Enriched |
| **6** | YWHAE | 7.79E-04 | 1.39 | 1.39 | 4 | 5 | Leak-Enriched |
| **7** | KRT1 | 1.57E-03 | 1.39 | 2.70 | 4 | 4 | Leak-Enriched |
| **8** | PNLIP | 7.41E-06 | 3.06 | 4.37 | 3 | 5 | Leak-Enriched |
| **9** | PRSS2 | 1.06E-02 | 3.01 | 3.23 | 3 | 5 | Leak-Enriched |
| **10** | CPB1 | 5.11E-04 | 2.96 | 3.53 | 3 | 5 | Leak-Enriched |
| **11** | CPA1 | 3.41E-06 | 2.91 | 4.80 | 3 | 5 | Leak-Enriched |
| **12** | AMY2A | 1.74E-07 | 2.75 | 4.96 | 3 | 5 | Leak-Enriched |
| **13** | IGLV7-46 | 5.12E-03 | 2.05 | 2.48 | 3 | 5 | Leak-Enriched |
| **14** | REG1A | 3.32E-02 | 1.99 | 2.37 | 3 | 5 | Leak-Enriched |
| **15** | CELA2A | 1.68E-02 | 1.98 | 2.94 | 3 | 5 | Leak-Enriched |
| **16** | EPX | 5.15E-03 | -1.82 | -2.26 | 3 | 5 | Non-Leak-Enriched |
| **17** | HBG1 | 1.02E-02 | 1.72 | 2.08 | 3 | 5 | Leak-Enriched |
| **18** | MMP9 | 1.57E-03 | 1.61 | 2.45 | 3 | 5 | Leak-Enriched |
| **19** | LXN | 8.21E-04 | 1.56 | 1.92 | 3 | 5 | Leak-Enriched |
| **20** | CAT | 7.96E-03 | 1.40 | 1.60 | 3 | 5 | Leak-Enriched |
| **21** | CD47 | 6.40E-02 | 1.29 | 1.58 | 3 | 5 | Leak-Enriched |
| **22** | HBD | 5.12E-03 | 1.22 | 1.95 | 3 | 5 | Leak-Enriched |
| **23** | FCGR3B | 1.70E-03 | 1.21 | 1.25 | 3 | 5 | Leak-Enriched |
| **24** | HPRT1 | 1.75E-02 | 1.16 | 1.26 | 3 | 5 | Leak-Enriched |
| **25** | PSMA3 | 5.15E-03 | 1.15 | 1.30 | 3 | 5 | Leak-Enriched |
| **26** | PNP | 2.48E-02 | 1.15 | 1.39 | 3 | 5 | Leak-Enriched |
| **27** | RPS10 | 2.08E-04 | -1.13 | -1.42 | 3 | 5 | Non-Leak-Enriched |
| **28** | NME1 | 1.15E-02 | 1.13 | 1.26 | 3 | 5 | Leak-Enriched |
| **29** | SOD1 | 2.35E-03 | 1.13 | 1.72 | 3 | 5 | Leak-Enriched |
| **30** | PSMB2 | 8.99E-04 | 1.07 | 1.19 | 3 | 5 | Leak-Enriched |
| **31** | CA1 | 1.00E-02 | 1.04 | 1.51 | 3 | 5 | Leak-Enriched |
| **32** | BPGM | 1.23E-02 | 1.03 | 1.52 | 3 | 5 | Leak-Enriched |
| **33** | GIMAP4 | 9.15E-03 | -1.02 | -1.03 | 3 | 5 | Non-Leak-Enriched |
| **34** | SELENBP1 | 1.53E-02 | 1.01 | 1.32 | 3 | 5 | Leak-Enriched |
| **35** | AZU1 | 1.69E-03 | 1.95 | 2.48 | 3 | 4 | Leak-Enriched |
| **36** | IGHV3OR16-13 | 7.65E-03 | 1.74 | 2.58 | 3 | 4 | Leak-Enriched |
| **37** | LTF | 1.98E-03 | 1.68 | 2.34 | 3 | 4 | Leak-Enriched |
| **38** | MPO | 7.28E-03 | 1.61 | 2.08 | 3 | 4 | Leak-Enriched |
| **39** | H2BC12 | 1.91E-04 | 1.37 | 2.39 | 3 | 4 | Leak-Enriched |
| **40** | CEACAM8 | 9.15E-03 | 1.09 | 2.27 | 3 | 4 | Leak-Enriched |
| **41** | AMY2B | 1.28E-07 | 3.34 | 5.38 | 2 | 5 | Leak-Enriched |
| **42** | PRSS1 | 2.60E-05 | 2.78 | 4.34 | 2 | 5 | Leak-Enriched |
| **43** | HBG2 | 5.50E-02 | 1.66 | 2.26 | 2 | 5 | Leak-Enriched |
| **44** | PFN1 | 1.10E-03 | 1.55 | 3.00 | 2 | 5 | Leak-Enriched |
| **45** | EVPL | 7.15E-02 | -1.15 | -2.04 | 2 | 5 | Non-Leak-Enriched |
| **46** | GPI | 1.32E-02 | 1.11 | 1.48 | 2 | 5 | Leak-Enriched |
| **47** | CAMP | 1.10E-03 | 1.06 | 1.75 | 2 | 5 | Leak-Enriched |
| **48** | S100A6 | 1.17E-02 | 1.03 | 1.62 | 2 | 5 | Leak-Enriched |
| **49** | KRT2 | 1.21E-02 | 1.02 | 1.95 | 2 | 5 | Leak-Enriched |
| **50** | IGKV2-40 | 8.13E-03 | -1.00 | -1.24 | 2 | 5 | Non-Leak-Enriched |
| **51** | KRT10 | 2.50E-03 | 0.99 | 2.27 | 2 | 5 | Leak-Enriched |
| **52** | PRDX6 | 7.91E-03 | 0.97 | 1.45 | 2 | 5 | Leak-Enriched |
| **53** | CSF1R | 2.27E-03 | -0.97 | -1.11 | 2 | 5 | Non-Leak-Enriched |
| **54** | TKT | 4.32E-03 | 0.96 | 1.69 | 2 | 5 | Leak-Enriched |
| **55** | LGALS3 | 1.10E-03 | 0.94 | 1.48 | 2 | 5 | Leak-Enriched |
| **56** | HPR | 5.49E-02 | -0.94 | -1.05 | 2 | 5 | Non-Leak-Enriched |
| **57** | S100A11 | 2.82E-03 | 0.94 | 1.41 | 2 | 5 | Leak-Enriched |
| **58** | PITHD1 | 1.00E-02 | 0.94 | 1.34 | 2 | 5 | Leak-Enriched |
| **59** | PGK1 | 2.10E-03 | 0.94 | 1.39 | 2 | 5 | Leak-Enriched |
| **60** | TALDO1 | 6.16E-03 | 0.91 | 1.38 | 2 | 5 | Leak-Enriched |
| **61** | PRDX2 | 4.00E-02 | 0.90 | 1.47 | 2 | 5 | Leak-Enriched |
| **62** | SLC4A1 | 5.99E-02 | 0.90 | 1.80 | 2 | 5 | Leak-Enriched |
| **63** | PARK7 | 1.09E-02 | 0.89 | 1.10 | 2 | 5 | Leak-Enriched |
| **64** | MYH9 | 8.80E-03 | 0.88 | 1.41 | 2 | 5 | Leak-Enriched |
| **65** | PYGL | 3.07E-03 | 0.87 | 1.43 | 2 | 5 | Leak-Enriched |
| **66** | ITIH3 | 4.32E-03 | -0.87 | -1.42 | 2 | 5 | Non-Leak-Enriched |
| **67** | PROCR | 3.02E-03 | -0.85 | -1.29 | 2 | 5 | Non-Leak-Enriched |
| **68** | PCMT1 | 1.53E-02 | 0.85 | 1.22 | 2 | 5 | Leak-Enriched |
| **69** | BLVRB | 2.02E-02 | 0.84 | 1.65 | 2 | 5 | Leak-Enriched |
| **70** | MST1 | 1.96E-03 | -0.84 | -1.37 | 2 | 5 | Non-Leak-Enriched |
| **71** | RAN | 3.25E-02 | 0.82 | 1.19 | 2 | 5 | Leak-Enriched |
| **72** | ARPC3 | 3.44E-02 | 0.82 | 1.13 | 2 | 5 | Leak-Enriched |
| **73** | PSMA5 | 6.85E-03 | 0.80 | 1.16 | 2 | 5 | Leak-Enriched |
| **74** | IGKC | 7.01E-04 | -0.79 | -1.08 | 2 | 5 | Non-Leak-Enriched |
| **75** | UBE2V1 | 1.64E-03 | 0.78 | 1.56 | 2 | 5 | Leak-Enriched |
| **76** | IGKV1-39 | 1.40E-02 | -0.75 | -1.36 | 2 | 5 | Non-Leak-Enriched |
| **77** | RAB1A | 8.03E-03 | 0.74 | 1.13 | 2 | 5 | Leak-Enriched |
| **78** | MYL9 | 5.59E-03 | 0.73 | 1.63 | 2 | 5 | Leak-Enriched |
| **79** | PGD | 1.45E-02 | 0.73 | 1.25 | 2 | 5 | Leak-Enriched |
| **80** | CSTB | 5.14E-03 | 0.72 | 1.12 | 2 | 5 | Leak-Enriched |
| **81** | APOB | 3.31E-02 | -0.67 | -1.21 | 2 | 5 | Non-Leak-Enriched |
| **82** | ARG1 | 5.12E-03 | 0.67 | 1.36 | 2 | 5 | Leak-Enriched |
| **83** | TBCA | 1.59E-04 | 0.66 | 1.61 | 2 | 5 | Leak-Enriched |
| **84** | ST13 | 5.57E-04 | 0.65 | 1.54 | 2 | 5 | Leak-Enriched |
| **85** | ARPC2 | 5.57E-03 | 0.63 | 1.09 | 2 | 5 | Leak-Enriched |
| **86** | ACTN4 | 7.06E-03 | 0.60 | 1.06 | 2 | 5 | Leak-Enriched |
| **87** | CAPZA1 | 1.43E-02 | 0.57 | 1.02 | 2 | 5 | Leak-Enriched |
| **88** | TXN | 2.50E-03 | 0.54 | 1.09 | 2 | 5 | Leak-Enriched |
| **89** | CELA3A | 3.39E-02 | 1.72 | 3.02 | 2 | 4 | Leak-Enriched |
| **90** | LYZ | 1.10E-03 | 1.42 | 2.43 | 2 | 4 | Leak-Enriched |
| **91** | PRTN3 | 3.05E-03 | 1.40 | 2.04 | 2 | 4 | Leak-Enriched |
| **92** | S100A9 | 6.97E-03 | 1.36 | 1.55 | 2 | 4 | Leak-Enriched |
| **93** | S100A8 | 8.13E-03 | 1.33 | 1.44 | 2 | 4 | Leak-Enriched |
| **94** | KRT9 | 3.85E-02 | 1.23 | 1.96 | 2 | 4 | Leak-Enriched |
| **95** | LCN2 | 5.14E-03 | 1.23 | 1.70 | 2 | 4 | Leak-Enriched |
| **96** | H3-7 | 4.77E-04 | 1.20 | 2.24 | 2 | 4 | Leak-Enriched |
| **97** | H2BC26 | 6.64E-02 | 1.16 | 1.46 | 2 | 4 | Leak-Enriched |
| **98** | ANXA3 | 2.70E-03 | 1.11 | 2.54 | 2 | 4 | Leak-Enriched |
| **99** | SERPINB1 | 1.28E-02 | 1.09 | 1.54 | 2 | 4 | Leak-Enriched |
| **100** | ACTN2 | 9.25E-02 | 0.87 | 1.36 | 2 | 4 | Leak-Enriched |
| **101** | CTSG | 4.15E-03 | 0.97 | 1.94 | 2 | 4 | Leak-Enriched |
| **102** | ACTA1 | 1.56E-02 | 0.53 | 1.09 | 2 | 4 | Leak-Enriched |
| **103** | H4C16 | 1.10E-03 | 0.93 | 2.06 | 2 | 4 | Leak-Enriched |
| **104** | ACTN1 | 1.15E-02 | 0.91 | 1.36 | 2 | 4 | Leak-Enriched |
| **105** | MMP8 | 6.06E-03 | 0.91 | 1.85 | 2 | 4 | Leak-Enriched |
| **106** | S100A12 | 2.77E-03 | 0.60 | -1.20 | 2 | 4 | Leak-Enriched |
| **107** | HSP90AA1 | 5.43E-02 | 0.81 | 1.61 | 2 | 4 | Leak-Enriched |
| **108** | LCP1 | 3.05E-03 | 0.80 | 1.27 | 2 | 4 | Leak-Enriched |
| **109** | ARPC5 | 2.46E-02 | 0.78 | 1.18 | 2 | 4 | Leak-Enriched |
| **110** | BPI | 6.89E-03 | 0.72 | 2.00 | 2 | 4 | Leak-Enriched |
| **111** | PGLYRP1 | 5.48E-02 | 0.62 | 1.09 | 2 | 4 | Leak-Enriched |
| **112** | FOLR3 | 8.03E-02 | 2.36 | 3.60 | 1 | 5 | Leak-Enriched |
| **113** | CELA3B | 4.86E-02 | 2.23 | 2.84 | 1 | 5 | Leak-Enriched |
| **114** | GP2 | 4.93E-02 | 2.20 | 3.67 | 1 | 5 | Leak-Enriched |
| **115** | CTRB2 | 4.12E-02 | 1.93 | 2.10 | 1 | 5 | Leak-Enriched |
| **116** | TMSB4X | 4.23E-02 | 1.54 | 2.09 | 1 | 5 | Leak-Enriched |
| **117** | PFDN6 | 4.69E-02 | 1.16 | 1.48 | 1 | 5 | Leak-Enriched |
| **118** | APOA4 | 2.41E-02 | -1.13 | -1.17 | 1 | 5 | Non-Leak-Enriched |
| **119** | IGHV3-35 | 9.44E-02 | 1.12 | 1.26 | 1 | 5 | Leak-Enriched |
| **120** | TSNAX | 5.30E-02 | 1.03 | 1.58 | 1 | 5 | Leak-Enriched |
| **121** | MYLK | 5.26E-02 | 1.02 | 1.15 | 1 | 5 | Leak-Enriched |
| **122** | SOD2 | 1.07E-02 | 1.00 | 1.29 | 1 | 5 | Leak-Enriched |
| **123** | PRG2 | 1.75E-02 | -0.98 | -1.51 | 1 | 5 | Non-Leak-Enriched |
| **124** | SPTA1 | 3.72E-02 | 0.96 | 1.55 | 1 | 5 | Leak-Enriched |
| **125** | IGKV1-16 | 7.16E-02 | -0.94 | -1.16 | 1 | 5 | Non-Leak-Enriched |
| **126** | MDH1 | 1.68E-02 | 0.93 | 1.11 | 1 | 5 | Leak-Enriched |
| **127** | SPTB | 6.58E-02 | 0.93 | 1.45 | 1 | 5 | Leak-Enriched |
| **128** | HAGH | 3.06E-02 | 0.92 | 1.06 | 1 | 5 | Leak-Enriched |
| **129** | OLFM4 | 4.47E-02 | 0.90 | 1.72 | 1 | 5 | Leak-Enriched |
| **130** | STOM | 1.77E-02 | 0.89 | 1.80 | 1 | 5 | Leak-Enriched |
| **131** | ALAD | 4.14E-02 | 0.88 | 1.41 | 1 | 5 | Leak-Enriched |
| **132** | RPIA | 5.41E-02 | 0.88 | 1.23 | 1 | 5 | Leak-Enriched |
| **133** | LTA4H | 5.57E-03 | 0.87 | 1.14 | 1 | 5 | Leak-Enriched |
| **134** | CA2 | 3.06E-02 | 0.86 | 1.45 | 1 | 5 | Leak-Enriched |
| **135** | LDHA | 1.34E-02 | 0.86 | 1.15 | 1 | 5 | Leak-Enriched |
| **136** | TNC | 1.28E-03 | -0.86 | -1.02 | 1 | 5 | Non-Leak-Enriched |
| **137** | PTCD1 | 4.59E-02 | -0.85 | -1.22 | 1 | 5 | Non-Leak-Enriched |
| **138** | TPM3 | 1.55E-02 | 0.85 | 1.39 | 1 | 5 | Leak-Enriched |
| **139** | IGHG4 | 5.48E-02 | -0.84 | -1.12 | 1 | 5 | Non-Leak-Enriched |
| **140** | PSMF1 | 4.57E-02 | 0.84 | 1.30 | 1 | 5 | Leak-Enriched |
| **141** | ANK1 | 5.88E-02 | 0.84 | 1.49 | 1 | 5 | Leak-Enriched |
| **142** | DSC2 | 1.89E-02 | -0.84 | -1.11 | 1 | 5 | Non-Leak-Enriched |
| **143** | FTCD | 6.90E-02 | 0.83 | 1.69 | 1 | 5 | Leak-Enriched |
| **144** | IDH1 | 1.19E-02 | 0.82 | 1.09 | 1 | 5 | Leak-Enriched |
| **145** | FUS | 6.62E-02 | -0.82 | -1.34 | 1 | 5 | Non-Leak-Enriched |
| **146** | PROS1 | 2.72E-02 | 0.82 | 1.21 | 1 | 5 | Leak-Enriched |
| **147** | GLO1 | 1.88E-02 | 0.80 | 1.02 | 1 | 5 | Leak-Enriched |
| **148** | ADSL | 2.95E-02 | 0.79 | 1.29 | 1 | 5 | Leak-Enriched |
| **149** | GAPDH | 9.15E-03 | 0.78 | 1.16 | 1 | 5 | Leak-Enriched |
| **150** | IGKV1D-8 | 2.44E-03 | -0.77 | -1.33 | 1 | 5 | Non-Leak-Enriched |
| **151** | IGHV1-46 | 1.13E-02 | -0.77 | -1.42 | 1 | 5 | Non-Leak-Enriched |
| **152** | EPB41 | 3.78E-02 | 0.76 | 1.38 | 1 | 5 | Leak-Enriched |
| **153** | PSMB1 | 1.89E-02 | 0.75 | 1.04 | 1 | 5 | Leak-Enriched |
| **154** | CLPS | 7.24E-02 | 0.74 | 1.57 | 1 | 5 | Leak-Enriched |
| **155** | TPI1 | 3.43E-02 | 0.74 | 1.11 | 1 | 5 | Leak-Enriched |
| **156** | MFAP4 | 6.10E-02 | -0.74 | -1.37 | 1 | 5 | Non-Leak-Enriched |
| **157** | TSN | 2.87E-02 | 0.73 | 1.25 | 1 | 5 | Leak-Enriched |
| **158** | GKN1 | 8.13E-03 | -0.72 | -1.64 | 1 | 5 | Non-Leak-Enriched |
| **159** | APEH | 1.00E-02 | 0.72 | 1.35 | 1 | 5 | Leak-Enriched |
| **160** | PTPRF | 1.10E-03 | -0.72 | -1.16 | 1 | 5 | Non-Leak-Enriched |
| **161** | SHBG | 3.01E-02 | -0.71 | -1.07 | 1 | 5 | Non-Leak-Enriched |
| **162** | IGLL1 | 2.48E-02 | -0.69 | -1.06 | 1 | 5 | Non-Leak-Enriched |
| **163** | UBE2N | 1.85E-02 | 0.68 | 1.17 | 1 | 5 | Leak-Enriched |
| **164** | COL18A1 | 3.61E-02 | -0.68 | -1.01 | 1 | 5 | Non-Leak-Enriched |
| **165** | ALDH1A1 | 1.62E-03 | 0.68 | 1.18 | 1 | 5 | Leak-Enriched |
| **166** | PRDX1 | 5.72E-04 | 0.67 | 1.59 | 1 | 5 | Leak-Enriched |
| **167** | IGHV1-18 | 6.46E-02 | -0.67 | -1.02 | 1 | 5 | Non-Leak-Enriched |
| **168** | ADH1B | 5.06E-02 | 0.67 | 1.11 | 1 | 5 | Leak-Enriched |
| **169** | IGHV5-51 | 1.17E-02 | -0.66 | -1.13 | 1 | 5 | Non-Leak-Enriched |
| **170** | ENO1 | 7.44E-03 | 0.66 | 1.23 | 1 | 5 | Leak-Enriched |
| **171** | PSMA1 | 2.24E-02 | 0.66 | 1.03 | 1 | 5 | Leak-Enriched |
| **172** | MYDGF | 1.09E-02 | 0.66 | 1.22 | 1 | 5 | Leak-Enriched |
| **173** | PPIB | 3.80E-03 | 0.65 | 1.01 | 1 | 5 | Leak-Enriched |
| **174** | CNTN1 | 1.45E-02 | -0.65 | -1.33 | 1 | 5 | Non-Leak-Enriched |
| **175** | HMGB1 | 2.82E-03 | 0.64 | 1.41 | 1 | 5 | Leak-Enriched |
| **176** | UBE2D3 | 3.13E-02 | 0.62 | 1.25 | 1 | 5 | Leak-Enriched |
| **177** | STX7 | 1.66E-02 | 0.61 | 1.24 | 1 | 5 | Leak-Enriched |
| **178** | TXNDC17 | 1.00E-02 | 0.60 | 1.05 | 1 | 5 | Leak-Enriched |
| **179** | HBA2 | 5.62E-02 | 0.60 | 2.20 | 1 | 5 | Leak-Enriched |
| **180** | C1QB | 9.27E-03 | -0.60 | -1.16 | 1 | 5 | Non-Leak-Enriched |
| **181** | MYL12A | 2.50E-03 | 0.60 | 1.56 | 1 | 5 | Leak-Enriched |
| **182** | BLVRA | 5.83E-03 | 0.58 | 1.19 | 1 | 5 | Leak-Enriched |
| **183** | UBE2L3 | 2.98E-02 | 0.58 | 1.09 | 1 | 5 | Leak-Enriched |
| **184** | C8G | 3.85E-02 | -0.58 | -1.04 | 1 | 5 | Non-Leak-Enriched |
| **185** | UROD | 1.10E-02 | 0.57 | 1.16 | 1 | 5 | Leak-Enriched |
| **186** | PSME1 | 5.36E-04 | 0.57 | 1.06 | 1 | 5 | Leak-Enriched |
| **187** | SPP2 | 3.85E-02 | -0.56 | -1.62 | 1 | 5 | Non-Leak-Enriched |
| **188** | ARHGEF6 | 6.85E-03 | -0.55 | -2.17 | 1 | 5 | Non-Leak-Enriched |
| **189** | EPB42 | 7.59E-02 | 0.55 | 1.32 | 1 | 5 | Leak-Enriched |
| **190** | C4A | 9.37E-02 | 0.54 | 1.14 | 1 | 5 | Leak-Enriched |
| **191** | GPX1 | 1.88E-02 | 0.53 | 1.33 | 1 | 5 | Leak-Enriched |
| **192** | PTPA | 4.78E-03 | 0.53 | 1.23 | 1 | 5 | Leak-Enriched |
| **193** | ADD1 | 3.13E-02 | 0.52 | 1.14 | 1 | 5 | Leak-Enriched |
| **194** | EEF1A1 | 5.35E-02 | 0.51 | 1.16 | 1 | 5 | Leak-Enriched |
| **195** | PSMB5 | 2.02E-02 | 0.51 | 1.04 | 1 | 5 | Leak-Enriched |
| **196** | UCHL5 | 6.30E-02 | 0.49 | 1.44 | 1 | 5 | Leak-Enriched |
| **197** | CP | 3.85E-02 | -0.47 | -1.01 | 1 | 5 | Non-Leak-Enriched |
| **198** | AK1 | 2.02E-02 | 0.47 | 1.39 | 1 | 5 | Leak-Enriched |
| **199** | ITIH4 | 5.54E-02 | -0.45 | -1.02 | 1 | 5 | Non-Leak-Enriched |
| **200** | ASPH | 1.57E-02 | -0.45 | -1.08 | 1 | 5 | Non-Leak-Enriched |
| **201** | TGFB1 | 8.26E-02 | 0.44 | 1.14 | 1 | 5 | Leak-Enriched |
| **202** | IGHG3 | 3.74E-02 | -0.44 | -1.02 | 1 | 5 | Non-Leak-Enriched |
| **203** | FETUB | 1.94E-02 | -0.41 | -1.09 | 1 | 5 | Non-Leak-Enriched |
| **204** | WDR77 | 2.71E-03 | 0.41 | 1.04 | 1 | 5 | Leak-Enriched |
| **205** | LGALS1 | 5.46E-02 | 0.37 | 1.04 | 1 | 5 | Leak-Enriched |
| **206** | ADGRF5 | 5.17E-02 | -0.35 | -1.06 | 1 | 5 | Non-Leak-Enriched |
| **207** | FABP4 | 9.01E-02 | 0.35 | 1.25 | 1 | 5 | Leak-Enriched |
| **208** | FBXO7 | 6.50E-02 | 0.35 | 1.04 | 1 | 5 | Leak-Enriched |
| **209** | PGM3 | 3.17E-03 | 0.28 | 1.03 | 1 | 5 | Leak-Enriched |
| **210** | PPBP | 4.14E-02 | 0.25 | 1.42 | 1 | 5 | Leak-Enriched |
| **211** | PSAT1 | 6.83E-02 | 0.25 | 1.05 | 1 | 5 | Leak-Enriched |
| **212** | COPS6 | 3.62E-02 | 0.17 | 1.10 | 1 | 5 | Leak-Enriched |
| **213** | IGHV3-38 | 1.58E-02 | -0.13 | -1.30 | 1 | 5 | Non-Leak-Enriched |
| **214** | CTRC | 7.03E-02 | 1.14 | 2.81 | 1 | 4 | Leak-Enriched |
| **215** | MACROH2A1 | 2.24E-02 | 0.96 | 1.58 | 1 | 4 | Leak-Enriched |
| **216** | HNRNPC | 3.44E-03 | -0.94 | -1.23 | 1 | 4 | Non-Leak-Enriched |
| **217** | CRP | 2.06E-02 | 0.78 | 1.37 | 1 | 4 | Leak-Enriched |
| **218** | SLC2A1 | 7.19E-02 | 0.92 | 1.55 | 1 | 4 | Leak-Enriched |
| **219** | PDXP | 4.57E-02 | 0.90 | 2.50 | 1 | 4 | Leak-Enriched |
| **220** | ELANE | 4.39E-02 | 0.89 | 1.39 | 1 | 4 | Leak-Enriched |
| **221** | RETN | 1.53E-02 | 0.43 | 1.42 | 1 | 4 | Leak-Enriched |
| **222** | DEFA3 | 4.73E-02 | 0.78 | 1.14 | 1 | 4 | Leak-Enriched |
| **223** | RNASE1 | 9.39E-04 | -0.78 | -1.33 | 1 | 4 | Non-Leak-Enriched |
| **224** | ITGB2 | 3.33E-03 | 0.71 | 1.63 | 1 | 4 | Leak-Enriched |
| **225** | PGLYRP2 | 4.52E-02 | -0.71 | -1.07 | 1 | 4 | Non-Leak-Enriched |
| **226** | GCA | 5.53E-02 | 0.69 | 1.51 | 1 | 4 | Leak-Enriched |
| **227** | HK3 | 1.59E-02 | 0.67 | 1.04 | 1 | 4 | Leak-Enriched |
| **228** | PNPO | 8.73E-02 | 0.36 | 1.21 | 1 | 4 | Leak-Enriched |
| **229** | DMTN | 5.37E-02 | 0.65 | 1.33 | 1 | 4 | Leak-Enriched |
| **230** | HRNR | 3.54E-02 | 0.54 | 1.48 | 1 | 4 | Leak-Enriched |
| **231** | PSMB7 | 3.52E-02 | 0.62 | 1.01 | 1 | 4 | Leak-Enriched |
| **232** | RAP2B | 8.18E-02 | 0.61 | 1.04 | 1 | 4 | Leak-Enriched |
| **233** | ARPC4 | 4.59E-02 | 0.60 | 1.01 | 1 | 4 | Leak-Enriched |
| **234** | FCN1 | 8.70E-02 | 0.59 | 1.15 | 1 | 4 | Leak-Enriched |
| **235** | IGLV3-10 | 8.72E-02 | 0.58 | 1.21 | 1 | 4 | Leak-Enriched |
| **236** | RAD23A | 4.57E-02 | 0.57 | 1.16 | 1 | 4 | Leak-Enriched |
| **237** | ITGAM | 1.77E-02 | 0.56 | 1.37 | 1 | 4 | Leak-Enriched |
| **238** | GNB2 | 4.86E-02 | 0.56 | 1.49 | 1 | 4 | Leak-Enriched |
| **239** | DKK3 | 2.11E-02 | -0.15 | -1.01 | 1 | 4 | Non-Leak-Enriched |
| **240** | RANBP1 | 4.82E-02 | 0.52 | 1.09 | 1 | 4 | Leak-Enriched |
| **241** | EFHD2 | 8.13E-03 | 0.50 | 1.17 | 1 | 4 | Leak-Enriched |
| **242** | ASAP1 | 9.51E-02 | 0.47 | 1.04 | 1 | 4 | Leak-Enriched |
| **243** | ACTR2 | 2.46E-02 | 0.47 | 1.11 | 1 | 4 | Leak-Enriched |
| **244** | ISLR | 9.25E-02 | -0.46 | -1.27 | 1 | 4 | Non-Leak-Enriched |
| **245** | ESD | 3.69E-02 | 0.44 | 1.13 | 1 | 4 | Leak-Enriched |
| **246** | ANXA1 | 7.31E-02 | 0.44 | 1.15 | 1 | 4 | Leak-Enriched |
| **247** | KRT18 | 3.34E-02 | 0.43 | 1.00 | 1 | 4 | Leak-Enriched |
| **248** | PSME3 | 6.21E-02 | -0.27 | -2.68 | 1 | 4 | Non-Leak-Enriched |
| **249** | CNRIP1 | 5.22E-02 | 0.37 | 1.17 | 1 | 4 | Leak-Enriched |
| **250** | SEC14L4 | 8.28E-02 | 0.36 | 1.19 | 1 | 4 | Leak-Enriched |
| **251** | RAD23B | 6.09E-02 | 0.31 | 1.17 | 1 | 4 | Leak-Enriched |
| **252** | CBR1 | 1.64E-02 | 0.11 | 1.05 | 1 | 4 | Leak-Enriched |
| **253** | RHOA | 3.93E-02 | 0.19 | 1.05 | 1 | 4 | Leak-Enriched |
| **254** | HDDC2 | 1.02E-02 | 0.17 | 1.34 | 1 | 4 | Leak-Enriched |
| **255** | TIMP1 | 1.02E-02 | -0.12 | -1.88 | 1 | 4 | Non-Leak-Enriched |
| **256** | LSM3 | 7.24E-02 | -0.09 | 1.41 | 1 | 4 | Non-Leak-Enriched |

**Supplementary Table S2. Full ranked list of differentially expressed proteins in the colorectal cohort across postoperative days.**

| **Rank** | **Protein** | **BestPerPOD_FDR** | **MedLog2FC** | **PeakLog2FC** | **Sign. POD’s** | **Same Sign POD’s** | **Direction** |
| --- | --- | --- | --- | --- | --- | --- | --- |
| 1 | BTD | 6.40E-05 | -1.34 | -1.64 | 4 | 5 | Non-Leak-Enriched |
| 2 | CFHR1 | 1.39E-02 | -1.03 | -1.38 | 3 | 5 | Non-Leak-Enriched |
| 3 | HBG1 | 3.73E-02 | 1.70 | 2.61 | 2 | 5 | Leak-Enriched |
| 4 | TMOD1 | 1.58E-02 | 1.48 | 1.93 | 2 | 5 | Leak-Enriched |
| 5 | TPM1 | 1.56E-02 | -1.30 | -1.97 | 2 | 5 | Non-Leak-Enriched |
| 6 | RHCE | 3.33E-02 | 1.28 | 1.75 | 2 | 5 | Leak-Enriched |
| 7 | GMPR | 1.57E-02 | 1.28 | 2.17 | 2 | 5 | Leak-Enriched |
| 8 | TPM3 | 1.38E-03 | -1.27 | -1.33 | 2 | 5 | Non-Leak-Enriched |
| 9 | UBAC1 | 7.21E-02 | 1.22 | 1.34 | 2 | 5 | Leak-Enriched |
| 10 | AHSG | 4.82E-04 | -1.15 | -1.92 | 2 | 5 | Non-Leak-Enriched |
| 11 | SPP2 | 3.82E-02 | -1.12 | -1.49 | 2 | 5 | Non-Leak-Enriched |
| 12 | CPN2 | 3.53E-04 | -1.12 | -1.41 | 2 | 5 | Non-Leak-Enriched |
| 13 | STOM | 2.26E-02 | 1.12 | 1.68 | 2 | 5 | Leak-Enriched |
| 14 | PPP2R5D | 1.57E-02 | 1.02 | 1.66 | 2 | 5 | Leak-Enriched |
| 15 | CDH5 | 5.21E-03 | -1.00 | -1.26 | 2 | 5 | Non-Leak-Enriched |
| 16 | MPP1 | 4.37E-02 | 1.00 | 1.06 | 2 | 5 | Leak-Enriched |
| 17 | PAFAH1B2 | 2.42E-02 | 0.99 | 1.20 | 2 | 5 | Leak-Enriched |
| 18 | RPIA | 2.17E-02 | 0.98 | 1.11 | 2 | 5 | Leak-Enriched |
| 19 | GPS1 | 2.69E-02 | 0.97 | 1.16 | 2 | 5 | Leak-Enriched |
| 20 | PI16 | 8.62E-04 | -0.96 | -1.26 | 2 | 5 | Non-Leak-Enriched |
| 21 | ROBO4 | 1.56E-02 | -0.95 | -1.22 | 2 | 5 | Non-Leak-Enriched |
| 22 | CNDP1 | 3.53E-04 | -0.93 | -1.63 | 2 | 5 | Non-Leak-Enriched |
| 23 | CBR3 | 3.81E-02 | 0.92 | 1.34 | 2 | 5 | Leak-Enriched |
| 24 | TKFC | 1.19E-02 | 0.91 | 1.27 | 2 | 5 | Leak-Enriched |
| 25 | DDI2 | 9.12E-03 | 0.88 | 1.60 | 2 | 5 | Leak-Enriched |
| 26 | PKLR | 1.57E-02 | 0.87 | 1.36 | 2 | 5 | Leak-Enriched |
| 27 | SERPINA6 | 9.58E-04 | -0.84 | -1.38 | 2 | 5 | Non-Leak-Enriched |
| 28 | PON1 | 1.27E-02 | -0.76 | -1.40 | 2 | 5 | Non-Leak-Enriched |
| 29 | C4BPB | 8.62E-04 | -0.75 | -1.30 | 2 | 5 | Non-Leak-Enriched |
| 30 | LZIC | 4.91E-02 | 0.74 | 1.45 | 2 | 5 | Leak-Enriched |
| 31 | UBE2V1 | 2.40E-02 | 0.72 | 1.30 | 2 | 5 | Leak-Enriched |
| 32 | RAB5B | 4.82E-04 | 0.68 | 1.94 | 2 | 5 | Leak-Enriched |
| 33 | PROZ | 9.12E-03 | -0.66 | -1.48 | 2 | 5 | Non-Leak-Enriched |
| 34 | GPLD1 | 1.63E-03 | -0.62 | -1.26 | 2 | 5 | Non-Leak-Enriched |
| 35 | UROD | 3.81E-02 | 0.62 | 1.33 | 2 | 5 | Leak-Enriched |
| 36 | F12 | 9.00E-03 | -0.53 | -1.05 | 2 | 5 | Non-Leak-Enriched |
| 37 | HMBS | 2.76E-02 | 0.45 | 1.21 | 2 | 5 | Leak-Enriched |
| 38 | MYADM | 4.81E-02 | 0.30 | 1.51 | 2 | 5 | Leak-Enriched |
| 39 | EIF5 | 6.11E-02 | 0.50 | 2.28 | 2 | 4 | Leak-Enriched |
| 40 | APOA4 | 1.42E-02 | -1.23 | -1.61 | 2 | 4 | Non-Leak-Enriched |
| 41 | CPNE3 | 4.82E-04 | 1.19 | 2.02 | 2 | 4 | Leak-Enriched |
| 42 | CEACAM6 | 6.48E-02 | 1.16 | 3.45 | 2 | 4 | Leak-Enriched |
| 43 | HBZ | 5.67E-02 | 1.15 | 1.43 | 2 | 4 | Leak-Enriched |
| 44 | CEACAM8 | 1.88E-02 | 0.69 | 2.27 | 2 | 4 | Leak-Enriched |
| 45 | ANXA3 | 6.61E-02 | 0.45 | 1.56 | 2 | 4 | Leak-Enriched |
| 46 | RAB27A | 5.44E-02 | 0.40 | 1.46 | 2 | 4 | Leak-Enriched |
| 47 | ARL6IP5 | 3.23E-02 | 0.27 | 2.19 | 2 | 4 | Leak-Enriched |
| 48 | RHAG | 8.73E-02 | 1.68 | 2.21 | 1 | 5 | Leak-Enriched |
| 49 | IGLV3-12 | 8.85E-02 | -1.48 | -1.92 | 1 | 5 | Non-Leak-Enriched |
| 50 | IGLV3-10 | 6.43E-02 | -1.37 | -2.87 | 1 | 5 | Non-Leak-Enriched |
| 51 | SPARC | 1.70E-02 | -1.27 | -2.11 | 1 | 5 | Non-Leak-Enriched |
| 52 | SNCA | 8.45E-02 | 1.26 | 1.49 | 1 | 5 | Leak-Enriched |
| 53 | NCAM1 | 8.02E-02 | -1.15 | -2.23 | 1 | 5 | Non-Leak-Enriched |
| 54 | CAT | 4.49E-02 | 1.10 | 1.15 | 1 | 5 | Leak-Enriched |
| 55 | FLOT2 | 5.43E-02 | 1.00 | 1.07 | 1 | 5 | Leak-Enriched |
| 56 | NIPSNAP3A | 6.47E-02 | 0.99 | 1.98 | 1 | 5 | Leak-Enriched |
| 57 | SH3BGRL | 1.52E-02 | 0.99 | 1.15 | 1 | 5 | Leak-Enriched |
| 58 | PSMG1 | 5.61E-02 | 0.99 | 1.71 | 1 | 5 | Leak-Enriched |
| 59 | HDHD2 | 5.00E-02 | 0.96 | 1.07 | 1 | 5 | Leak-Enriched |
| 60 | PPP2R1A | 2.10E-02 | 0.95 | 1.87 | 1 | 5 | Leak-Enriched |
| 61 | PRPSAP1 | 4.16E-02 | 0.94 | 1.76 | 1 | 5 | Leak-Enriched |
| 62 | SERPINA3 | 1.25E-02 | -0.93 | -1.21 | 1 | 5 | Non-Leak-Enriched |
| 63 | CFH | 2.93E-03 | -0.91 | -1.72 | 1 | 5 | Non-Leak-Enriched |
| 64 | PZP | 7.90E-02 | -0.91 | -1.15 | 1 | 5 | Non-Leak-Enriched |
| 65 | SPTB | 6.51E-02 | 0.90 | 1.41 | 1 | 5 | Leak-Enriched |
| 66 | NAPA | 3.12E-02 | 0.90 | 1.13 | 1 | 5 | Leak-Enriched |
| 67 | FN3K | 1.97E-02 | 0.90 | 1.70 | 1 | 5 | Leak-Enriched |
| 68 | FBXO7 | 1.57E-02 | 0.88 | 1.80 | 1 | 5 | Leak-Enriched |
| 69 | AGO2 | 7.15E-02 | 0.87 | 1.06 | 1 | 5 | Leak-Enriched |
| 70 | HPRT1 | 7.53E-03 | 0.86 | 1.39 | 1 | 5 | Leak-Enriched |
| 71 | CD44 | 1.63E-03 | -0.86 | -1.22 | 1 | 5 | Non-Leak-Enriched |
| 72 | ACYP1 | 4.91E-02 | 0.84 | 1.55 | 1 | 5 | Leak-Enriched |
| 73 | NIF3L1 | 1.58E-02 | 0.84 | 1.36 | 1 | 5 | Leak-Enriched |
| 74 | FGR | 7.09E-02 | 0.84 | 1.49 | 1 | 5 | Leak-Enriched |
| 75 | GCLM | 7.15E-02 | 0.83 | 1.10 | 1 | 5 | Leak-Enriched |
| 76 | HPR | 6.01E-02 | -0.82 | -1.34 | 1 | 5 | Non-Leak-Enriched |
| 77 | DMTN | 1.57E-02 | 0.81 | 1.84 | 1 | 5 | Leak-Enriched |
| 78 | FH | 4.41E-02 | 0.81 | 1.17 | 1 | 5 | Leak-Enriched |
| 79 | PGM2L1 | 2.07E-02 | 0.79 | 1.29 | 1 | 5 | Leak-Enriched |
| 80 | UBE2N | 1.71E-02 | 0.79 | 1.21 | 1 | 5 | Leak-Enriched |
| 81 | RAB13 | 6.57E-02 | 0.79 | 1.10 | 1 | 5 | Leak-Enriched |
| 82 | IPO9 | 7.14E-02 | 0.78 | 3.02 | 1 | 5 | Leak-Enriched |
| 83 | PRPS1 | 7.16E-02 | 0.78 | 1.35 | 1 | 5 | Leak-Enriched |
| 84 | PSMB3 | 5.20E-02 | 0.78 | 1.09 | 1 | 5 | Leak-Enriched |
| 85 | PRKCD | 4.65E-02 | 0.77 | 1.21 | 1 | 5 | Leak-Enriched |
| 86 | SPTA1 | 7.15E-02 | 0.77 | 1.40 | 1 | 5 | Leak-Enriched |
| 87 | UBE2L3 | 2.21E-02 | 0.75 | 1.08 | 1 | 5 | Leak-Enriched |
| 88 | CP | 2.70E-03 | -0.74 | -1.02 | 1 | 5 | Non-Leak-Enriched |
| 89 | ACOT7 | 7.15E-02 | 0.74 | 1.29 | 1 | 5 | Leak-Enriched |
| 90 | NUTF2 | 5.25E-02 | 0.74 | 1.24 | 1 | 5 | Leak-Enriched |
| 91 | VCAM1 | 2.09E-03 | -0.73 | -1.02 | 1 | 5 | Non-Leak-Enriched |
| 92 | COPS7B | 2.76E-02 | 0.72 | 1.30 | 1 | 5 | Leak-Enriched |
| 93 | BASP1 | 4.56E-02 | -0.72 | -1.19 | 1 | 5 | Non-Leak-Enriched |
| 94 | EIF1B | 4.29E-02 | 0.72 | 1.57 | 1 | 5 | Leak-Enriched |
| 95 | CFHR2 | 1.39E-02 | -0.69 | -1.11 | 1 | 5 | Non-Leak-Enriched |
| 96 | CLSTN1 | 3.73E-02 | -0.68 | -1.09 | 1 | 5 | Non-Leak-Enriched |
| 97 | ADSL | 2.04E-02 | 0.68 | 1.02 | 1 | 5 | Leak-Enriched |
| 98 | GCLC | 9.62E-02 | 0.68 | 1.05 | 1 | 5 | Leak-Enriched |
| 99 | C20orf27 | 1.66E-02 | 0.68 | 1.21 | 1 | 5 | Leak-Enriched |
| 100 | AKR7A2 | 4.36E-03 | 0.68 | 1.14 | 1 | 5 | Leak-Enriched |
| 101 | PPBP | 4.62E-02 | 0.67 | 1.31 | 1 | 5 | Leak-Enriched |
| 102 | IGHV1-24 | 5.00E-02 | 0.66 | 1.29 | 1 | 5 | Leak-Enriched |
| 103 | GFUS | 4.63E-02 | 0.65 | 1.34 | 1 | 5 | Leak-Enriched |
| 104 | SKP1 | 3.15E-02 | 0.65 | 1.30 | 1 | 5 | Leak-Enriched |
| 105 | CYB5R3 | 1.26E-02 | 0.63 | 1.05 | 1 | 5 | Leak-Enriched |
| 106 | FBLN1 | 5.21E-03 | -0.62 | -1.02 | 1 | 5 | Non-Leak-Enriched |
| 107 | PITHD1 | 3.95E-02 | 0.62 | 1.18 | 1 | 5 | Leak-Enriched |
| 108 | PAFAH1B3 | 4.47E-02 | 0.62 | 1.08 | 1 | 5 | Leak-Enriched |
| 109 | FABP4 | 7.57E-02 | -0.61 | -1.02 | 1 | 5 | Non-Leak-Enriched |
| 110 | CZIB | 6.09E-02 | 0.61 | 1.30 | 1 | 5 | Leak-Enriched |
| 111 | IGHV5-10-1 | 5.26E-02 | 0.61 | 1.10 | 1 | 5 | Leak-Enriched |
| 112 | SERPINF2 | 1.54E-02 | -0.60 | -1.03 | 1 | 5 | Non-Leak-Enriched |
| 113 | HBQ1 | 2.97E-02 | 0.60 | 1.41 | 1 | 5 | Leak-Enriched |
| 114 | CUL1 | 1.57E-02 | 0.59 | 1.16 | 1 | 5 | Leak-Enriched |
| 115 | MAP2K2 | 4.91E-03 | 0.59 | 1.31 | 1 | 5 | Leak-Enriched |
| 116 | USP9X | 4.99E-02 | 0.59 | 1.09 | 1 | 5 | Leak-Enriched |
| 117 | PADI2 | 2.16E-02 | 0.58 | 1.53 | 1 | 5 | Leak-Enriched |
| 118 | LGALS3BP | 9.00E-03 | -0.57 | -1.09 | 1 | 5 | Non-Leak-Enriched |
| 119 | ADD2 | 4.39E-02 | 0.56 | 1.38 | 1 | 5 | Leak-Enriched |
| 120 | AP2A2 | 3.98E-02 | 0.56 | 1.16 | 1 | 5 | Leak-Enriched |
| 121 | HRG | 2.57E-02 | -0.56 | -1.16 | 1 | 5 | Non-Leak-Enriched |
| 122 | UBE2I | 5.21E-03 | 0.55 | 1.09 | 1 | 5 | Leak-Enriched |
| 123 | PROS1 | 7.53E-03 | -0.55 | -1.03 | 1 | 5 | Non-Leak-Enriched |
| 124 | WDR77 | 4.21E-02 | 0.53 | 1.03 | 1 | 5 | Leak-Enriched |
| 125 | F9 | 7.53E-03 | -0.53 | -1.02 | 1 | 5 | Non-Leak-Enriched |
| 126 | ARG1 | 6.25E-02 | 0.53 | 1.35 | 1 | 5 | Leak-Enriched |
| 127 | HNRNPH1 | 4.29E-02 | 0.52 | 1.08 | 1 | 5 | Leak-Enriched |
| 128 | APOM | 4.61E-02 | -0.52 | -1.01 | 1 | 5 | Non-Leak-Enriched |
| 129 | ADK | 2.10E-02 | 0.51 | 1.08 | 1 | 5 | Leak-Enriched |
| 130 | DOK3 | 1.94E-02 | 0.49 | 1.32 | 1 | 5 | Leak-Enriched |
| 131 | NME2 | 2.72E-02 | 0.48 | 1.01 | 1 | 5 | Leak-Enriched |
| 132 | APOC3 | 8.10E-03 | -0.46 | -1.06 | 1 | 5 | Non-Leak-Enriched |
| 133 | BOLA2B | 6.93E-02 | 0.44 | 1.33 | 1 | 5 | Leak-Enriched |
| 134 | S100P | 1.57E-02 | 0.43 | 1.06 | 1 | 5 | Leak-Enriched |
| 135 | C4B_2 | 3.37E-02 | -0.43 | -1.00 | 1 | 5 | Non-Leak-Enriched |
| 136 | COPS5 | 1.57E-02 | 0.43 | 1.23 | 1 | 5 | Leak-Enriched |
| 137 | CAPNS1 | 3.43E-02 | 0.42 | 1.08 | 1 | 5 | Leak-Enriched |
| 138 | EIF4B | 7.01E-02 | 0.41 | 1.01 | 1 | 5 | Leak-Enriched |
| 139 | MOB1B | 1.63E-03 | 0.33 | 1.12 | 1 | 5 | Leak-Enriched |
| 140 | PCBP1 | 1.87E-02 | 0.31 | 1.15 | 1 | 5 | Leak-Enriched |
| 141 | PLBD1 | 3.73E-02 | 0.27 | 1.71 | 1 | 5 | Leak-Enriched |
| 142 | GRK2 | 8.99E-02 | 0.21 | 1.21 | 1 | 5 | Leak-Enriched |
| 143 | CCS | 5.98E-02 | 0.17 | 1.08 | 1 | 5 | Leak-Enriched |
| 144 | H2BC26 | 1.66E-02 | 1.54 | 1.56 | 1 | 4 | Leak-Enriched |
| 145 | IGHV8-51-1 | 1.87E-02 | 1.42 | 3.06 | 1 | 4 | Leak-Enriched |
| 146 | NAXE | 3.81E-02 | 1.29 | 1.42 | 1 | 4 | Leak-Enriched |
| 147 | PRPSAP2 | 2.41E-02 | 1.03 | 1.45 | 1 | 4 | Leak-Enriched |
| 148 | MOB3A | 9.28E-02 | 0.41 | -1.85 | 1 | 4 | Leak-Enriched |
| 149 | STOML3 | 3.82E-02 | 1.19 | 1.39 | 1 | 4 | Leak-Enriched |
| 150 | ACTR1B | 6.92E-02 | 1.17 | 2.01 | 1 | 4 | Leak-Enriched |
| 151 | SLC2A3 | 2.69E-02 | 1.13 | 1.42 | 1 | 4 | Leak-Enriched |
| 152 | CYP4F3 | 7.53E-03 | 1.10 | 2.47 | 1 | 4 | Leak-Enriched |
| 153 | SLC25A3 | 6.71E-02 | 1.10 | 1.35 | 1 | 4 | Leak-Enriched |
| 154 | CA4 | 5.36E-02 | 1.10 | 2.36 | 1 | 4 | Leak-Enriched |
| 155 | SH3GLB2 | 5.00E-02 | 0.82 | 1.32 | 1 | 4 | Leak-Enriched |
| 156 | AMY2A | 8.45E-02 | 1.02 | 3.29 | 1 | 4 | Leak-Enriched |
| 157 | RAD23A | 3.41E-02 | 1.02 | 1.32 | 1 | 4 | Leak-Enriched |
| 158 | IQGAP1 | 2.21E-02 | 1.01 | 1.07 | 1 | 4 | Leak-Enriched |
| 159 | SNRPA1 | 6.85E-02 | -0.93 | -1.24 | 1 | 4 | Non-Leak-Enriched |
| 160 | CTPS2 | 4.63E-02 | 0.99 | 1.31 | 1 | 4 | Leak-Enriched |
| 161 | ALOX5AP | 5.22E-02 | 0.97 | 1.69 | 1 | 4 | Leak-Enriched |
| 162 | LPCAT2 | 4.18E-02 | 0.91 | 1.55 | 1 | 4 | Leak-Enriched |
| 163 | NCF4 | 5.31E-02 | 0.91 | 1.09 | 1 | 4 | Leak-Enriched |
| 164 | SACM1L | 6.40E-02 | 0.90 | 1.12 | 1 | 4 | Leak-Enriched |
| 165 | MRI1 | 4.56E-02 | 0.89 | 1.23 | 1 | 4 | Leak-Enriched |
| 166 | CD47 | 7.53E-03 | 0.88 | 1.43 | 1 | 4 | Leak-Enriched |
| 167 | CUL2 | 5.00E-02 | 0.87 | 1.04 | 1 | 4 | Leak-Enriched |
| 168 | NUDCD2 | 8.46E-02 | 0.84 | 1.63 | 1 | 4 | Leak-Enriched |
| 169 | TPM4 | 2.97E-02 | -0.83 | -1.24 | 1 | 4 | Non-Leak-Enriched |
| 170 | PDXP | 7.15E-02 | 0.12 | 1.33 | 1 | 4 | Leak-Enriched |
| 171 | H1-0 | 3.23E-02 | 0.79 | 1.61 | 1 | 4 | Leak-Enriched |
| 172 | PPME1 | 1.97E-02 | 0.79 | 1.20 | 1 | 4 | Leak-Enriched |
| 173 | KRT4 | 9.35E-02 | -0.77 | -3.17 | 1 | 4 | Non-Leak-Enriched |
| 174 | MCEMP1 | 5.65E-03 | 0.77 | 2.35 | 1 | 4 | Leak-Enriched |
| 175 | ATP6V1D | 2.69E-02 | 0.76 | 1.84 | 1 | 4 | Leak-Enriched |
| 176 | RAP2A | 4.69E-02 | 0.72 | 1.44 | 1 | 4 | Leak-Enriched |
| 177 | PIP4K2A | 5.00E-02 | 0.71 | 1.14 | 1 | 4 | Leak-Enriched |
| 178 | PSMF1 | 3.15E-02 | 0.71 | 1.18 | 1 | 4 | Leak-Enriched |
| 179 | PPIL1 | 8.45E-02 | 0.71 | 1.05 | 1 | 4 | Leak-Enriched |
| 180 | GNG2 | 2.21E-02 | 0.70 | 1.38 | 1 | 4 | Leak-Enriched |
| 181 | RIC8A | 1.57E-02 | 0.66 | 1.72 | 1 | 4 | Leak-Enriched |
| 182 | CELA3B | 8.34E-02 | 0.63 | 2.94 | 1 | 4 | Leak-Enriched |
| 183 | COX4I1 | 8.34E-02 | 0.35 | -1.20 | 1 | 4 | Leak-Enriched |
| 184 | UFM1 | 1.26E-02 | 0.61 | 1.15 | 1 | 4 | Leak-Enriched |
| 185 | AHCYL1 | 7.50E-02 | -0.55 | -4.06 | 1 | 4 | Non-Leak-Enriched |
| 186 | VAPA | 5.22E-02 | 0.60 | 1.25 | 1 | 4 | Leak-Enriched |
| 187 | DTYMK | 5.16E-02 | 0.60 | 1.30 | 1 | 4 | Leak-Enriched |
| 188 | RILP | 7.15E-02 | 0.58 | 1.65 | 1 | 4 | Leak-Enriched |
| 189 | SCP2 | 5.11E-02 | 0.55 | 1.78 | 1 | 4 | Leak-Enriched |
| 190 | DNPH1 | 2.57E-02 | 0.53 | 1.56 | 1 | 4 | Leak-Enriched |
| 191 | RAP2B | 2.49E-02 | 0.53 | 1.01 | 1 | 4 | Leak-Enriched |
| 192 | CKM | 5.61E-02 | -0.51 | -1.02 | 1 | 4 | Non-Leak-Enriched |
| 193 | LSM6 | 9.71E-02 | 0.30 | -1.08 | 1 | 4 | Leak-Enriched |
| 194 | APEX1 | 4.91E-03 | 0.48 | 1.28 | 1 | 4 | Leak-Enriched |
| 195 | NBEAL2 | 7.09E-02 | 0.47 | 1.40 | 1 | 4 | Leak-Enriched |
| 196 | HMGB1 | 2.41E-02 | 0.29 | 1.22 | 1 | 4 | Leak-Enriched |
| 197 | CTSG | 6.33E-02 | 0.48 | 1.05 | 1 | 4 | Leak-Enriched |
| 198 | TOR1AIP1 | 2.63E-02 | 0.43 | 1.42 | 1 | 4 | Leak-Enriched |
| 199 | MAPRE1 | 8.62E-04 | 0.46 | 1.21 | 1 | 4 | Leak-Enriched |
| 200 | CS | 4.11E-02 | 0.23 | 1.58 | 1 | 4 | Leak-Enriched |
| 201 | NAP1L4 | 3.81E-02 | 0.35 | 1.08 | 1 | 4 | Leak-Enriched |
| 202 | CEACAM5 | 7.53E-03 | 0.42 | 1.83 | 1 | 4 | Leak-Enriched |
| 203 | CETP | 8.59E-02 | -0.40 | -1.11 | 1 | 4 | Non-Leak-Enriched |
| 204 | BPI | 6.91E-02 | 0.25 | 1.76 | 1 | 4 | Leak-Enriched |
| 205 | NCF1 | 3.62E-02 | 0.36 | 1.99 | 1 | 4 | Leak-Enriched |
| 206 | GSTM3 | 5.28E-02 | -0.36 | -2.44 | 1 | 4 | Non-Leak-Enriched |
| 207 | MACROH2A1 | 5.22E-02 | 0.34 | 1.29 | 1 | 4 | Leak-Enriched |
| 208 | DYSF | 3.37E-02 | 0.33 | 1.30 | 1 | 4 | Leak-Enriched |
| 209 | HUWE1 | 1.57E-02 | 0.32 | 1.59 | 1 | 4 | Leak-Enriched |
| 210 | PTBP3 | 9.47E-02 | 0.32 | 1.16 | 1 | 4 | Leak-Enriched |
| 211 | DPT | 4.62E-02 | -0.32 | -1.13 | 1 | 4 | Non-Leak-Enriched |
| 212 | S100A9 | 1.57E-02 | -0.24 | -1.26 | 1 | 4 | Non-Leak-Enriched |
| 213 | CYBB | 2.11E-02 | 0.19 | 1.49 | 1 | 4 | Leak-Enriched |
| 214 | FAM114A2 | 3.77E-02 | 0.06 | 1.39 | 1 | 4 | Leak-Enriched |

**Supplementary Table S3. Sample availability across postoperative days**

| **Cohort** | **POD** | **Non leak patients(n)** | **Leak patients (n)** |
| --- | --- | --- | --- |
| Colorectal | POD1 | 20 | 15 |
| Colorectal | POD2 | 20 | 15 |
| Colorectal | POD3 | 20 | 15 |
| Colorectal | POD4 | 16 | 8 |
| Colorectal | POD5 | 10 | 4 |
| Pancreatic | POD1 | 16 | 17 |
| Pancreatic | POD2 | 16 | 15 |
| Pancreatic | POD3 | 14 | 16 |
| Pancreatic | POD4 | 15 | 15 |
| Pancreatic | POD5 | 13 | 14 |
| Pancreatic | POD6 | 11 | 9 |
| Pancreatic | POD7 | 8 | 6 |
| Pancreatic | POD8 | 5 | 4 |
| Pancreatic | POD9 | 3 | 2 |
| Pancreatic | POD10 | 1 | 1 |
